# An epidemiological forecast model and software assessing interventions on COVID-19 epidemic in China

**DOI:** 10.1101/2020.02.29.20029421

**Authors:** Lili Wang, Yiwang Zhou, Jie He, Bin Zhu, Fei Wang, Lu Tang, Marisa Eisenberg, Peter X.K. Song

## Abstract

We develop a health informatics toolbox that enables public health workers to timely analyze and evaluate the time-course dynamics of the novel coronavirus (COVID-19) infection using the public available data from the China CDC. This toolbox is built upon a hierarchical epidemiological model in which two observed time series of daily proportions of infected and removed cases are emitted from the underlying infection dynamics governed by a Markov SIR infectious disease process. We extend the SIR model to incorporate various types of time-varying quarantine protocols, including government-level macro isolation policies and community-level micro inspection measures. We develop a calibration procedure for under-reported infected cases. This toolbox provides forecast, in both online and offline forms, of turning points of interest, including the time when daily infected proportion becomes smaller than the previous ones and the time when daily infected proportions becomes smaller than that of daily removed proportion, as well as the ending time of the epidemic. An R software is made available for the public, and examples on the use of this software are illustrated. Some possible extensions of our novel epidemiological models are discussed.

## 1 Introduction

The outbreak of the coronavirus disease or COVID-19, originated in Wuhan, the capital city of Hubei province, spreads out quickly in Hubei and then in China, South Korea, Japan, Iran and Italy, as well as many other cities across the world. Being one of the great epidemics in the 21st century, till February 25, 2020, in China it has caused a total of 78,195 confirmed infections, 2,718 deaths and 30,078 recovered cases, and 2,491 suspected cases still remained to be tested. Since the outbreak of the epidemic, many clinical papers [15, 3, 37, 38, 12, 6, 20, 5, 11, 8, 7, 28, 10, 42, 34] have been published to unveil limited but important knowledge of COVID-19, including that (i) COVID-19 is an infectious disease caused by SARS-CoV-2, a virus closely related to the SARS coronavirus (SARS-CoV) [20, 5, 32]; (ii) it can surely spread from person to person [11, 8]; (iii) it has a high person-to-person transmission rate, especially in the case with close contact, resulting in a large number of infected cases in Hubei province; (iv) the median incubation time is 3 days, which can be as long as 24 days [7]; and (v) asymptomatic person carrying SARS-CoV-2 is contagious [28]. This epidemic has been concerning not only in China but also in the rest of the world given the currently fast growing number of infected cases in South Korea, Japan, Iran, and Italy.

In addition to vaccination, quarantine or medical isolation is the human wisdom that has been proved to be one of the most effective ways to stop the spreading of infectious diseases such as SARS [35, 29, 22] and plague [4] in the human history. The basic idea of quarantine is to separate infected cases from susceptible population and *vice versa*. Since mid-January 2020, the Chinese government has implemented all kinds of very strict in-home isolation protocols nationwide, which have been elevated day by day through various government enforced quarantine and societally organized inspections to control the spread of COVID-19 in Hubei and other regions in China. In the meantime, the Chinese government has quickly increased the capacity of hospitals or as such that took symptomatic patients to be quarantined and treated by medical doctors and nurses, most of whom were dispatched from hospitals outside of Hubei province.

The question of the most importance, which draws most attention, concerns when the spread of COVID-19 will end. This question has to be answered by a prediction model using the daily most-updated data from the China CDC. Unfortunately, it is extremely difficult to make right and precise prediction due to the limited amount of available data, which are thought to be inaccurate due to the issue of under-reporting. Many prediction models [33, 18, 9, 27, 24, 40, 19] have already been proposed to provide good fitting results for the publicly available data that may be potentially under-reported. Each of these models may result in different predictions of turning points, such as the dates when the daily increased or the total number of infections begin to decrease. Since such forecasting needs to extrapolate a fitted model to a relatively distance future time after the last date with observed data, whichever the chosen model is used, the model itself will dictate prediction results. Many tuning methods have been developed by data scientists to tune a prediction model, producing only point predictions with no quantification of prediction uncertainty. In addition, data accuracy, in particular the issue of under-reporting, may cause bias in prediction, and ignoring this issue would lead to an optimistic prediction of early turning points. The issue of under-reporting may be attributed to the unsatisfactory sensitivity of the RNA test for SARS-CoV-2 or to the lack of enough kits for testing at the beginning of the outbreak among other logistic and political reasons. The Chinese government tried to correct these issues by using a new diagnostic protocol based on clinical symptoms starting at the first week of February. However, it undermines the quality of data collected in the early phase of the epidemic.

All the above points illustrate the complexity of human interventions on the spread of COVID-19, including but not limited to in-home quarantine, hospitalization, community enforcement of wearing masks, minimizing outdoor activities, and changed diagnostic criteria by the government. The prediction model must take such features into account in order to provide meaningful analyses and hopefully reasonable predictions. However, most existing prediction models do not have the capacity to incorporate changing interventions in reality, and with no such critical component of time-varying intervention in the model, predicted turning points would be untrustworthy. Our new model and analytic toolbox aim to fill in this significant gap.

We develop a health informatics toolbox, with an R package eSIR [1, 26], that helps accomplish the following specific aims:

AIM 1 : Utilize and calibrate publicly available data to understand the epidemiological trend of COVID-19 spread in Hubei province and the other regions of China.

AIM 2 : Incorporate time-varying quarantine protocols in the model of COVID-19 infection dynamics via an extension of the classical epidemiological SIR model. This dynamic infection system necessitates the forecast of the future trend of COVID-19 spread.

AIM 3 : Provide an R software package to health workers who can conveniently perform their own analyses using their substantive knowledge and perhaps better quality data from provinces in China or from other countries.

By no means the toolbox developed by us would give an accurate prediction of turning points, but rather we hope to provide a data analytic toolbox to people who may have better domain-specific knowledge and experience as well as high quality data to carry out independent predictions.

Our informatics toolbox is built upon a state-space model [41, 13, 31, 14] shown in Figure 1 with an extended Markov SIR model [16], which has the following key features: (i) Our model is specified with the temporally varying prevalence of susceptible, infected and removed (recovered and death) compartments, not directly on time series of respective counts; (ii) estimation and inference are carried out and implemented by the Markov Chain Monte Carlo (MCMC) algorithm; (iii) it outputs various sample draws from the posteriors of the model parameters (e.g. transmission and removal rates) and the underlying prevalence of susceptible, infected and removed compartments, as well as their credible intervals. The latter is of extreme importance to quantify prediction uncertainty. In addition, this toolbox provides predicted turning points, including (i) the date when daily increased number of infections begins to decrease or the time at which the second order derivative of the prevalence of infected compartment is zero (i.e. the turning point of infection acceleration to deceleration); and (ii) the date when daily number of removed cases is larger than that of infected cases, or the time at which the first derivative of the prevalence of infected compartment is zero (i.e. the turning point of zero infection speed). As a byproduct, the method also provides a predicted time when the COVID-19 epidemic ends.

**Figure 1:**
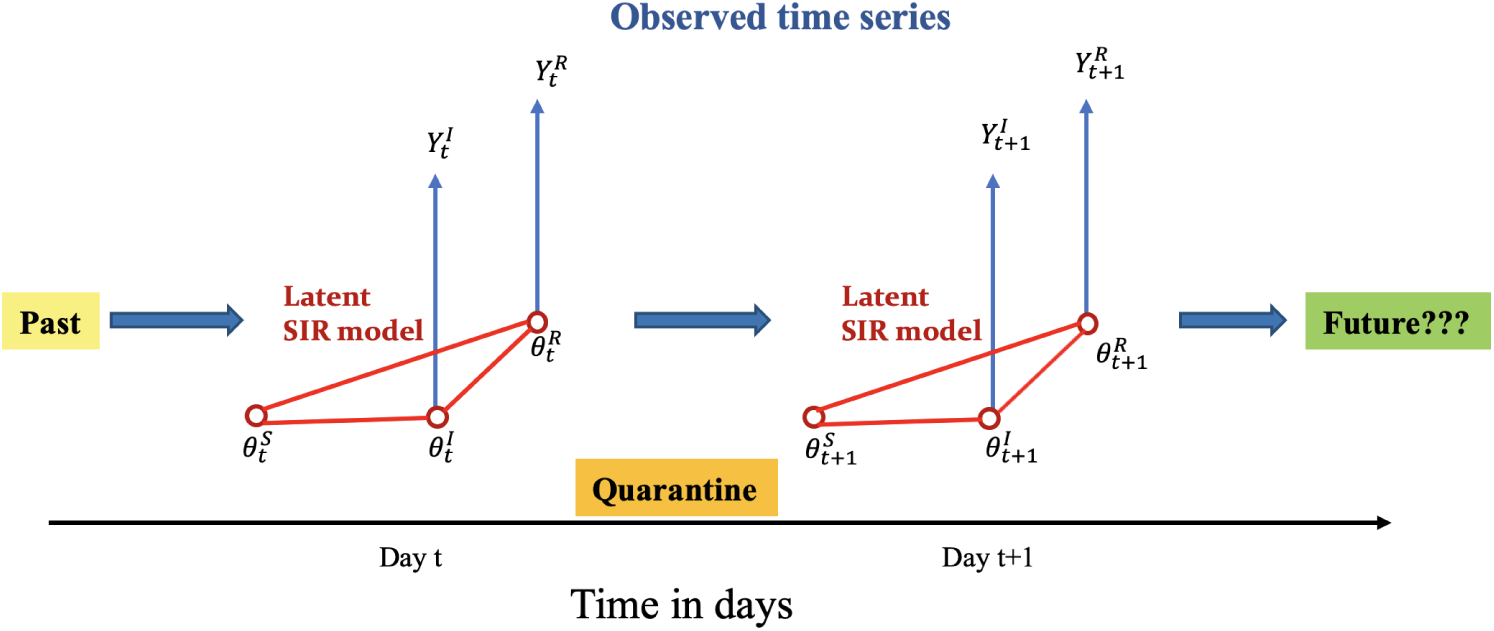
A conceptual framework of the proposed epidemiological state-space SIR model.

The R package `eSIR` is available at https://github.com/lilywang1988/eSIR. This paper is organized as follows. Section 2 presents our new epidemiological forecast model incorporating time-varying quarantine protocols. Section 3 concerns the algorithmic implementation via Markov Chain Monte Carlo and a deliverable R software. Section 4 is devoted to the analysis of COVID-19 data within and outside Hubei, where a calibration of under-reporting is proposed. Section 5 gives some concluding remarks, and some technical details are included in the appendices.

## 2 State-space SIR Epidemiological Model

### 2.1 Basic model of coronavirus infection

We begin with a basic epidemiological model in the framework of state-space SIR models with no consideration of quarantine protocols. This framework was proposed by [23] with only one-dimensional time series of infected proportions. Refer to Chapter 9-12 of [30] for an introduction to state-space models. Here we consider two time series of proportions of infected and removed cases, denoted by 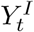 and 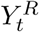 at time *t*, respectively, where the compartment of removed *R* is a sum of the proportions of recovered cases and deaths at time *t*. We assume that the 2-dimensional time series of 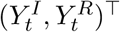 follows a state-space model with the beta distributions at time *t*:

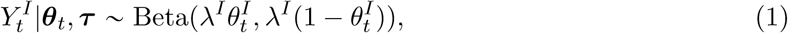

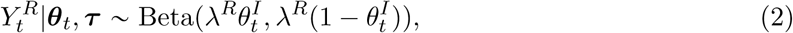

where 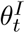 and 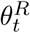 are the respective prevalence of infection and removal at time *t*, and *λ*^*I*^ and *λ*^*R*^ are the parameters controlling the respective variances of the observed proportions.

As shown in Figure 1, these observed time series are emitted from the underlying latent dynamics of COVID-19 infection characterized by the latent Markov process ***θ***_*t*_. It is easy to see that the expected proportions in both models (1) and (2) are equal to the prevalence of infection and the probability of removal at time *t*, namely 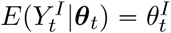 and 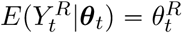. See Appendix B. Moreover, the latent population prevalence 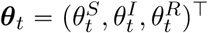 is a three-dimensional Markov process, in which 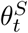 is the probability of a person being susceptible or at risk at time *t*, 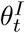 is the probability of a person being infected at time *t*, and 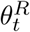 is the probability of a person being removed from the infectious system (either recovered or dead) at time *t*. Obviously, 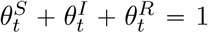. We assume that this 3-dimensional prevalence process ***θ***_*t*_ is governed by the following Markov model:

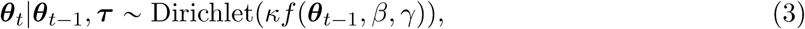

where parameter *κ* scales the variance of the Dirichlet distribution and function *f* (·) is a 3-dimensional vector that determines the mean of the Dirichlet distribution. The function *f* is the engine of the infection dynamics, which operates according to the classical infectious disease SIR model of the form:

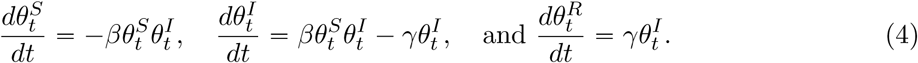

Note that the explicit solution to the above system (4) of ordinary differential equations is unavailable. Following [23], we invoke the fourth-order Runge–Kutta(RK4) approximation, resulting in an approximate of *f* (*θ*_*t*−_, *β, γ*) as follows:

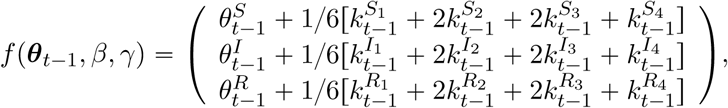

where all these *k*_*t*_ terms are given in the appendix A. Denote the set of model parameters by ***τ*** = (*β, γ*, ***θ***_0_, *λ, κ*)^T^, which will be estimated using the MCMC method [2].

### 2.2 Epidemiological model with time-varying transmission rate

The basic epidemiological model with both constant transmission and removal rates in SIR model (4) does not reflect the reality in China, where all levels of quarantines have been enforced. Many forms of human interventions that are altering the transmission rate over time include (i) individual-level protective measures such as wearing masks and safety glasses, using hygiene, and taking in-home isolation, and (ii) community-level quarantines such as hospitalization for infected cases, city blockade, traffic control and restricted social activities, and so on. In addition, the virus itself may mutate to evolve, so to increase the potential rate of false negative in the disease diagnosis. As a result, some individual virus carriers are not contained. Thus, the transmission rate *β* indeed varies over time, which should be accounted in the modeling.

One extension to the above basic epidemiological model is to allow a time-varying probability that a susceptible person meets an infected person or *vice versa*. Suppose at a time *t, q*^*S*^(*t*) ≈ [0, 1] is the chance of an at-risk person being in-home isolation, and *q*^*I*^(*t*) ≈ [0, 1] is the chance of an infected person being in-hospital quarantine. Thus, the chance of disease transmission when an at-risk person meets an infected person is modified as:

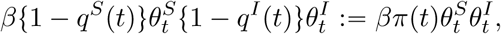

with *π*(*t*) := {1 − *q*^*S*^(*t*)}{1 − *q*^*I*^(*t*)} ∈ [0, 1]. In effect, this *π*(*t*) modifies the chance of a susceptible person meeting with an infected person or *vice versa*, which is termed as a *transmission modifier* due to quarantine in this paper. Obviously, with no quarantine in place, *π*(*t*) ≡ 1 for all time. See Figure 2. This results in a new SIR model with a time-varying transmission rate modifier:

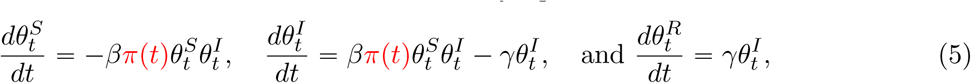

where the product term *βπ*(*t*) defines a realistic transmission rate reflective to the currently enforced quarantine measures of all levels in China. Note that the above extended SIR model assumes primarily that both population-level chance of being susceptible and population-level chance of being infected remain the same, but the chance of a susceptible person meeting with an infected person is reduced by *π*(*t*).

**Figure 2:**
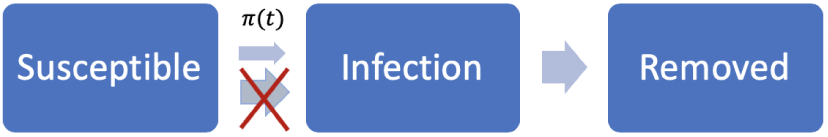
An extended SIR model with time-varying transmission rates.

The transmission rate modifier *π*(*t*) needs to be specified according to actual quarantine protocols in a given region. A possible choice of *π*(*t*) may be a step function that reflects government-initiated macro isolation measures in Wuhan, Hubei province:

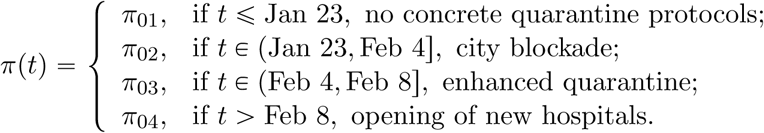

When ***π***_0_ = (*π*_01_, *π*_02_, *π*_03_, *π*_04_) are chosen with different values, as shown in Figure 3, we obtain different types of transmission rate modifiers aligned with different quarantine protocols.

**Figure 3:**
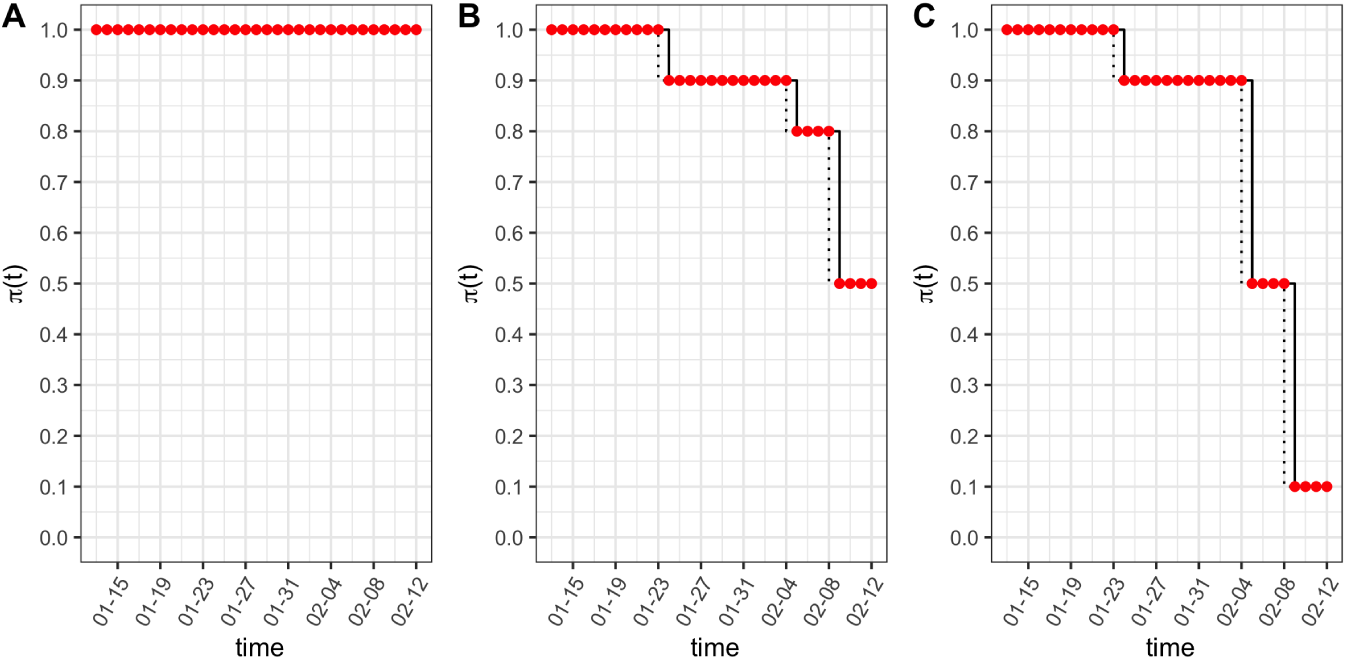
Transmission rate modifier takes different forms of step functions under different macro quarantine measures over time. Subfigures from A to C indicate ***π***_0_ = (*π*_01_, *π*_02_, *π*_03_, *π*_04_) equal to (1, 1, 1, 1), (1, 0.9, 0.8, 0.5) and (1, 0.9, 0.5, 0.1) with change time points at (Jan 23, Feb 4, Feb 8) respectively.

Alternatively, the modifier *π t* may be specified as a continuous function that reflects steadily increased community-level awareness and responsibility of voluntary quarantine and preventive measures, which may be regarded as a kind of micro isolation measure initiated by individuals or local self-organized inspections. For example, as shown in Figure 4, we may choose the following exponential functions:

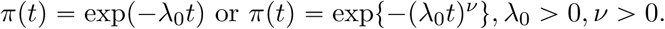

**Figure 4:**
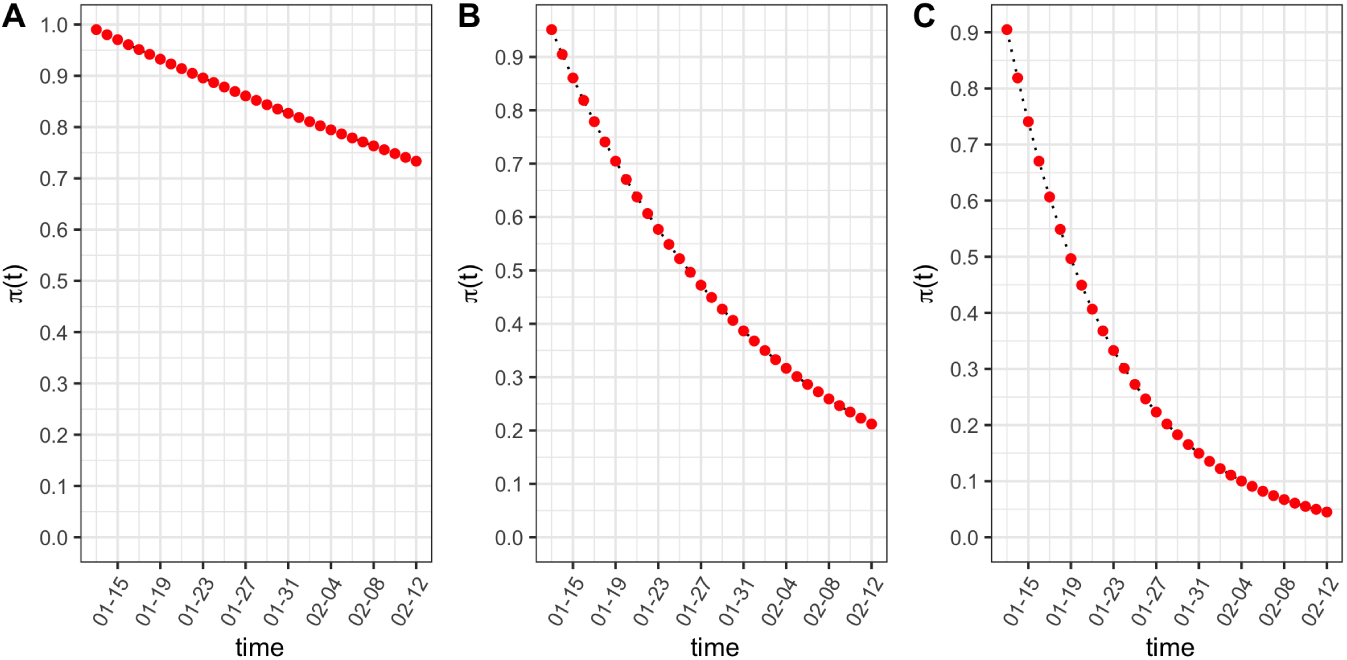
Transmission rate modifier takes continuous functions under difference micro quarantine measures over time. Subfigures from A to C are exponential survival functions with *λ*_0_ = 0.01,*λ*_0_ =0.05 and *λ*_0_ = 0.1 respectively.

Technically, the RK’s approximate of *f* function in Appendix A may be easily obtained by replacing *β* by *βπ*(*t*) in the specification of the latent prevalence model (3), and moreover in all quantities and steps in the MCMC implementation. See the detailed in Section 3.

### 2.3 Epidemiological model with quarantine compartment

An alternative way to incorporate quarantine measures into the basic epidemiological model (4) is to add a new quarantine compartment that collects quarantined individuals who would have no chance of meeting any infected individuals in the infection system, as shown in Figure 5. This model allows to characterize time-varying proportions of susceptible cases due largely to the government-enforced stringent in-home isolation outside of Hubei province. The basic SIR model in equation (4) is then extended by adding a quarantine compartment with a time-varying rate of quarantine *ϕ*(*t*), which is the chance of a susceptible person being willing to take in-home isolation at time *t*. The extended SIR takes the following 4-dimensional latent process 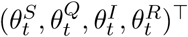:

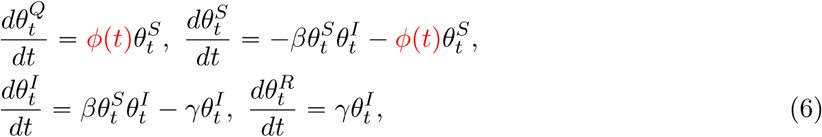

where 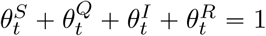.

**Figure 5:**
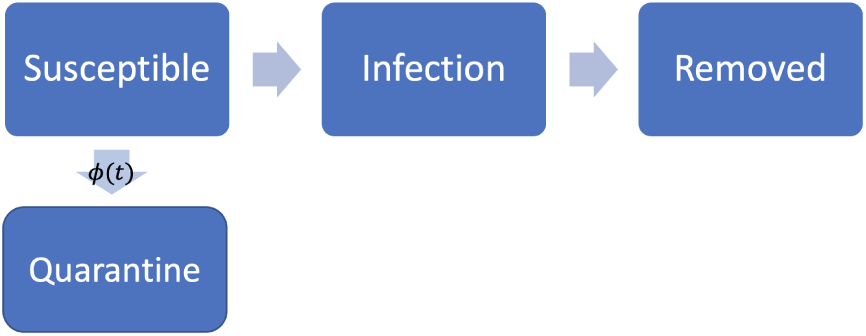
An extended SIR model with time-varying transmission rate.

We suppose that the quarantine rate *ϕ*(*t*) is a Dirac delta function with jumps at times when major macro quarantine measures are enforced. For example, we may specify the *ϕ*(*t*) function as follows:

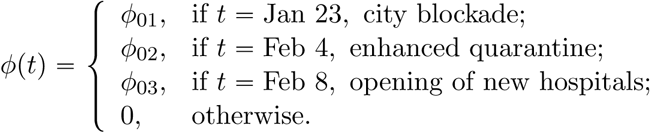

Here we show several examples of multi-point instantaneous quarantine rates in Figure 6 with jump sizes equal to ***ϕ***_0_ = (*ϕ*_01_, *ϕ*_02_, *ϕ*_03_) that occur respectively at dates of (Jan 23, Feb 4, Feb 8). In particular, we plot three scenarios, e.g., no intervention (***ϕ***_0_ = (0, 0, 0)), multiple moderate jumps (***ϕ***_0_ = (0.1, 0.4, 0.3)), and only one large jump (***ϕ***_0_ = (0, 0.9, 0)). Note that at each jump, the respective proportion of the susceptible population would move to the quarantine compartment. For example, with ***ϕ***_0_ = (0.1, 0.4, 0.3), the quarantine compartment will be enlarged accumulatively over three time points 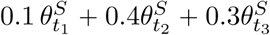.

**Figure 6:**
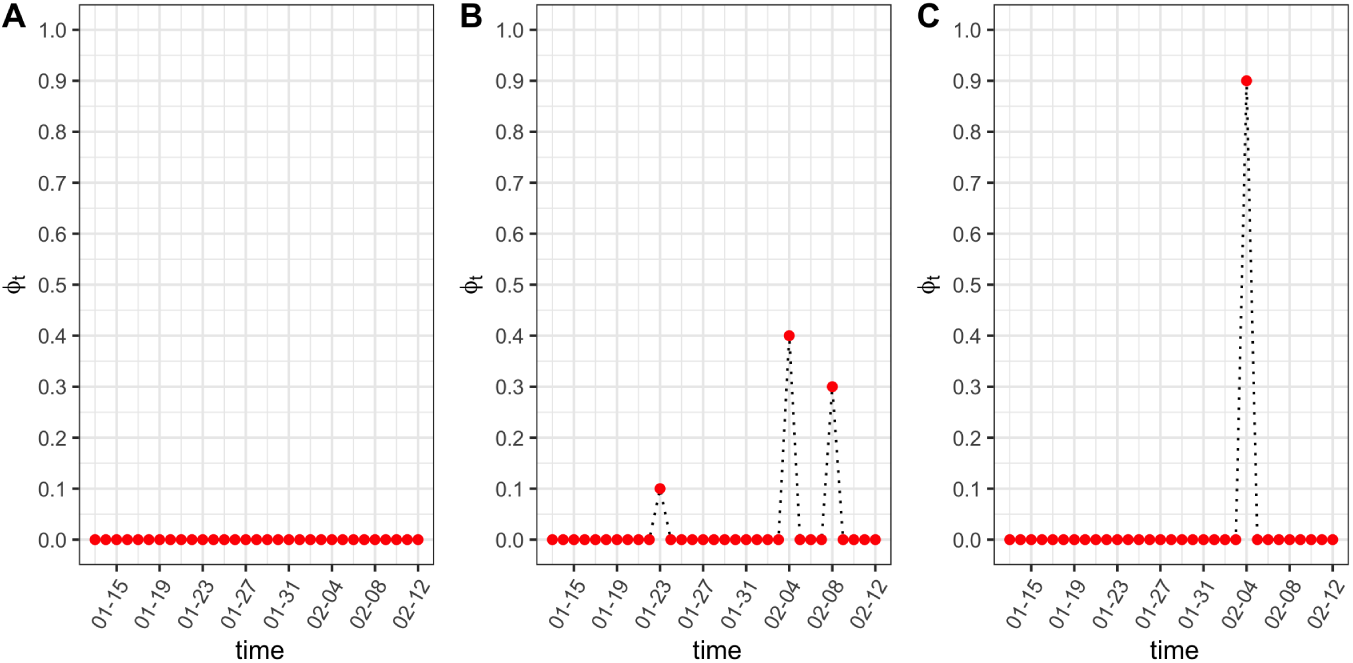
Three examples of multi-point instantaneous quarantine rates. Subfigures from A to C denote ***ϕ***_0_ = (0, 0, 0, 0), ***ϕ***_0_ = (0.1, 0.4, 0.3) and ***ϕ***_0_ = (0, 0.9, 0) at three time points of (Jan 23, Feb 4, Feb 8) respectively.

The *f* (***θ***_*t*−_, *β, γ*) function determined by the above extended SIR model (6) can be solved by applying the fourth-order Runge-Kutta approximation, and the resulting solution is given in

Appendix A. To deal with the Dirac delta function *ϕ*(*t*), we develop a two-step approximation for model (6). In brief, we first solve a continuous function without change points via the differential equations in (5), and then we directly move 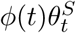 of the susceptible compartment to the quarantine compartment. From our experience, this approach largely improves the approximation accuracy in the presence of discontinuities.

## 3 Implementation: Markov Chain Monte Carlo Algorithm

### 3.1 MCMC Algorithm

We implemented the MCMC algorithm to collect draws from the posterior distributions, and further obtain posterior estimates and credible intervals of the unknown parameters in the above models specified in Section 2. Because of the hierarchical structure in the state-space model considered in this paper, the posterior distributions can be obtained straightforwardly. The R package rjags [25] can be directly applied to draw samples from the posterior distributions, so we omit the technical details. The latent Markov ***θ***_*t*_ processes are sampled and forecasted by the MCMC sampler, particularly for the prevalence of infection and the probability of removal, 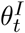 and 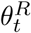, which enables us to determine turning points of interest and the reproduction number *R*_0_.

The first turning point of interest is the time when the daily number of new infected cases stops increasing. Mathematically, this corresponds to the time *t* at which 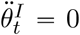 or the gradient of 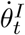 is zero. As illustrated by Panel A in Figure 7, the peak of 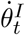, denoted by the vertical green line, is the first turning point of interest. The second turning point is the time when the cumulative infected cases reaches its maximum, meaning 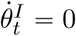. In principle, the second turning point occurs only after the first one, as shown in Panel B in Figure 7.

**Figure 7:**
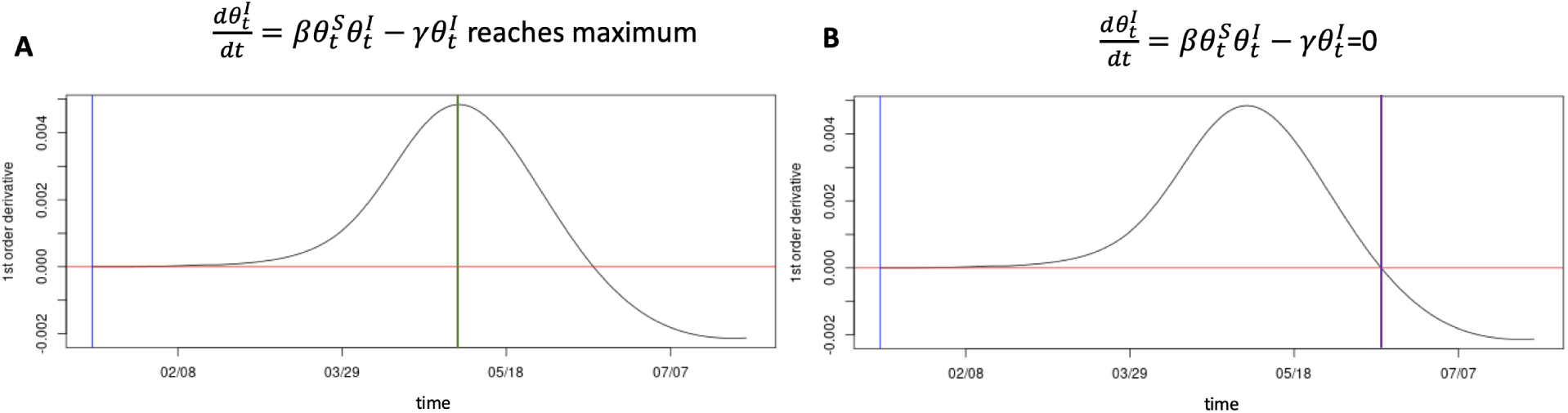
The first turning point in Panel A is marked by a green line, denoting the time when the estimated first-order derivative of the prevalence of infection reaches the maximum. The second turning point in Panel B is marked by a purple line, which is the time when the estimated first-order derivative of the prevalence of infection equals to zero. The vertical blue line labels the first observation day.

The reproduction number *R*_0_ of an infectious disease is estimated by the ratio *R*_0_ = *β*/*γ*, where *β* and *γ* are both estimated from their posterior distributions. Because our models consider the quarantine compartment, *R*_0_ might change according to the forms of quarantine protocols. We adopt a standard MCMC algorithm to draw the latent process. Let *t*_0_ be the current time up to which we have observed data 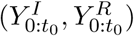. To perform *M* draws of 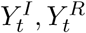 for *t* ∈ [*t*_0_ + 1, *T*], we proceed as follows: for each *m* = 1, …, *M*,

1. draw 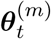 from the posterior 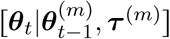 of the prevalence process, at *t* = *t*_0_ + 1, …, *T* ;
2. draw 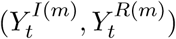 from 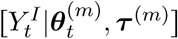 and 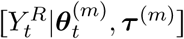 according to the observed process, at *t* = *t*_0_ + 1, …, *T*, respectively;

The prior distributions are specified with some of the hyper-parameters being set according to the SARS data from Hong Kong [21]. They are,

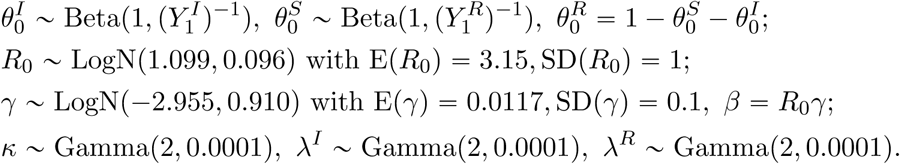

In the default setting the variances of the above prior distributions are set at relatively large values to reflect the fact that limited prior knowledge of these epidemiological model parameters is available. When more information becomes accessible during the course of the epidemic, smaller prior variance values may be used, leading to tighter credible intervals for the model parameters and turning points.

### 3.2 R software package

We deliver an R software package that is able to output the MCMC estimation, inference and prediction under the epidemiological model with two proposed extended SIR models that incorporate time-varying quarantine protocols. These new models have been discussed in detail in Sections 2.2 and 2.3. Our R package, named `eSIR`, uses daily-updated time series of infected and removed proportions as input data. The R package is available at GitHub `lilywang1988/eSIR`, and its user manual is appended as the supplementary material of this paper. The quarantine functions are predefined and will be treated as known functions of protocols/policies in the estimation and prediction steps. We let the transmission rate modifier *π*(*t*) be either a step function or an exponential function, and let the quarantine rate *ϕ*)*t*) follow a Dirac delta function with pre-specified points of jump and sizes of jumps. The package provides various plots for users to visualize the MCMC results, including the estimated prevalence of infection and the estimated probability of removal, and predicted turning points of interest. Various summary statistics are listed in the output, including posterior mean estimates of the transmission and removal rates, estimate of the reproduction number, and forecasts of turning points and their 95% credible intervals. Moreover, the package gives multiple options to users who can save the entire MCMC results, including the output tables and summary plots, traceplots for MCMC quality control, and full MCMC draws for user’s own summary analyses. Some illustrations on the use of this software package is given in Section 4. In addition, we developed an online R Shiny App that can automatically update the results whenever the China CDC updates the daily COVID-19 data.

## 4 Analysis of the COVID-19 Data Within and Outside Hubei

### 4.1 Calibration of under-reported infection data

Under-reporting of infections is a common issue in the data collection of infectious disease, especially at the beginning of an outbreak. When medical diagnostic tools become more accurate and reliable, as well the transparency of preventive measures gets improved for an exchange of voluntary in-home quarantine, certain adjustments in data typically occur. It is shown in Figure 8 that on Feb 12 the cumulative and daily added number of infected cases in Hubei had clear jumps with significantly large sizes. Such sizable jumps cannot happen within one day, rather they represent an accumulation of cases that have not been reported in previous dates prior to Feb 12. To fix this under-reporting issue, we develop a calibration procedure with the detail in Appendix C. Below we briefly describe our approach for the calibration of the infected cases.

**Figure 8:**
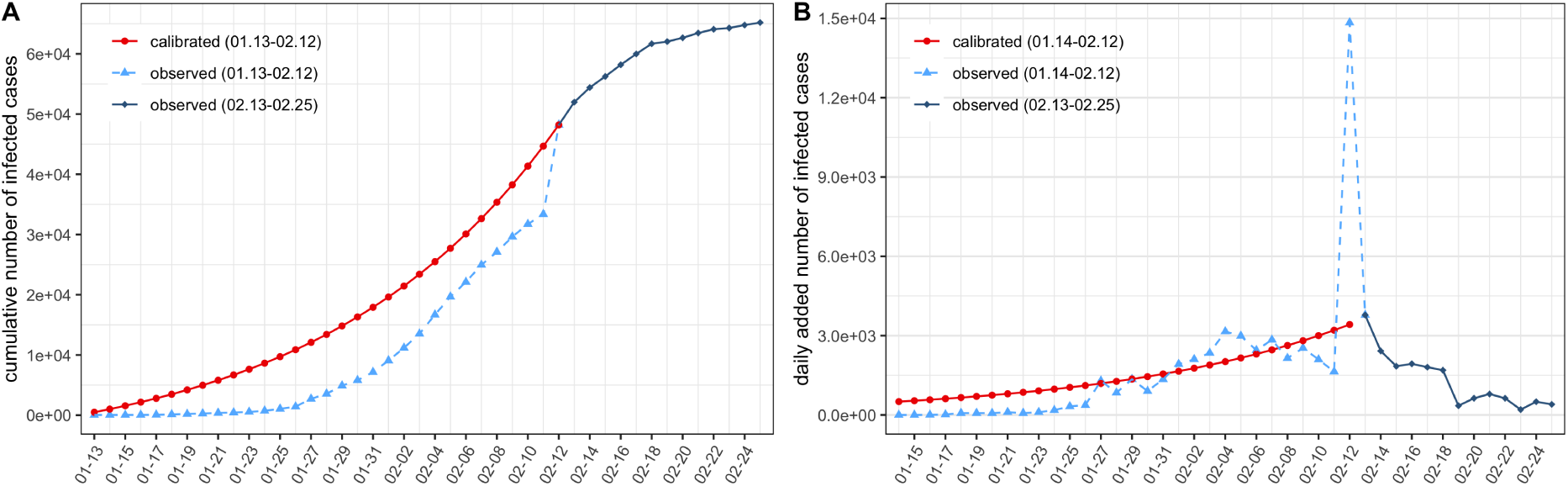
Under-reporting calibration of the infected cases in Hubei. (A) The cumulative number of infected cases. (B) The daily added number of infected cases. Data calibration is performed from Jan 13 to Feb 12 as is shown by the red curves.

We assume an exponential growth curve for the cumulative number of infected cases in Hubei before Feb 12 of the form *y*(*t*) = *ae*^*λt*^ + *b*, where parameters *λ, a, b* are to be estimated. Under the boundary conditions *y*(*t* = Jan 12) = 0 and *y*(*t* = Feb 12) = *a* exp(31*λ*) + *b*, we would like to minimize the one-step ahead extrapolation error on Feb 13. The constrained optimal solution can be obtained by the means of Lagrange Multipliers; the estimates are 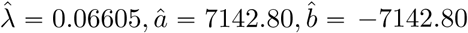. The resulting calibration curves for the cumulative and daily added number of infected cases are shown as the solid red curves in Figure 8. For example, on Jan 31, the reported cumulative number of infected cases is 7,153, but the calibrated number of infected cases is 17,911, with an increment of 10,758 cases. As shown later, this data quality control (QC) step helps improve the performance of MCMC.

### 4.2 Evaluation and prediction under time-varying quarantine

We applied our proposed models, algorithms and R package eSIR to analyze the COVID-19 data collected from the public website DXY.cn. The earliest public records for the provincial data are available since Jan 20, 2020. According to an existing R package on GitHub GuangchuangYu/nCov2019 [39], the total counts of confirmed infections and deaths are dated back on Jan 13, 2020. We assumed that before Jan 17 all the reported cases and deaths were from Hubei. We imputed by the linear interpolation the missing cases on Jan 18-19. Therefore, the data used in our analyses starts from Jan 13. The data used in analyses for the other provinces starts on Jan 23, which is the earliest time with non-zero values in the removed compartment. Note that there exist some minor discrepancies between different data sources, and the under-reporting issue is addressed in Appendix C by a calibration procedure. This section aims to provide a demonstration of our software to analyze the current public COVID-19 data, through which users may understand the proposed methods. We focus on illustrating ways to export and interpret the MCMC results. The R package may be applied to analyze infectious data from other countries.

First, we show the analysis of the Hubei COVID-19 data. Note that option `dic=T` enables to calculate the deviance information criterion (DIC) for model selection, while options, `save_files=T` and `save_mcmc`, allow the storage of MCMC output tables, plots, summary statistics and even full MCMC draws, which may be saved via the path of `file_add`, or otherwise via the current working directory. The major results returned from the package include predicted cumulative proportions, predicted turning points of interest, two ggplot2 [36] objects of the summary plots related to both infection and removed compartments, a summary output table containing all the posterior means, median and credible intervals of the model parameters, and DIC if opted. The traceplots and other useful diagnostic plots are also provided and automatically saved if `save_files=T` is opted. In the package, function `tvt.eSIR()` works on the epidemiological model with time-varying transmission rate in Section 2.2, and `qh.eSIR()` for the other epidemiological model with a quarantine compartment in Section 2.3. Note that in function `tvt.eSIR()`, with a choice of `exponential=F`, a step function is run in the MCMC when both `change_time` and `pi0` are specified. To fit the model with a continuous transmission rate modifier function, user may set `exponential=T` and specify a value of `lambda0`. The default is to run the basic epidemiological model with no quarantine or *π*(*t*) = 1 in Section 2.1. `death_in_R` is usually set to be the average ratio of death and removed proportions at each observation time point, which is used to estimate the death curve in the forecast plot of the removed compartment. Below are the R scripts used in the analysis.

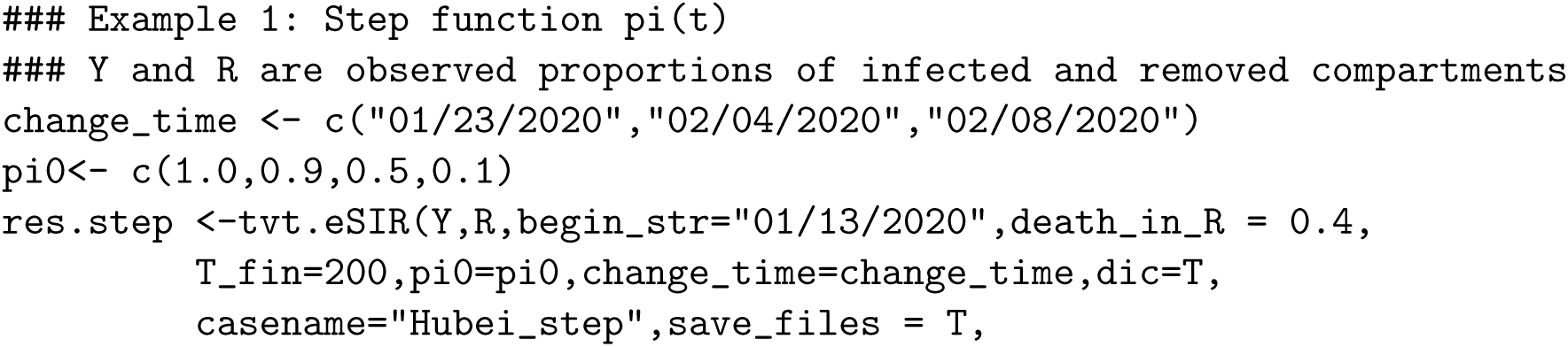
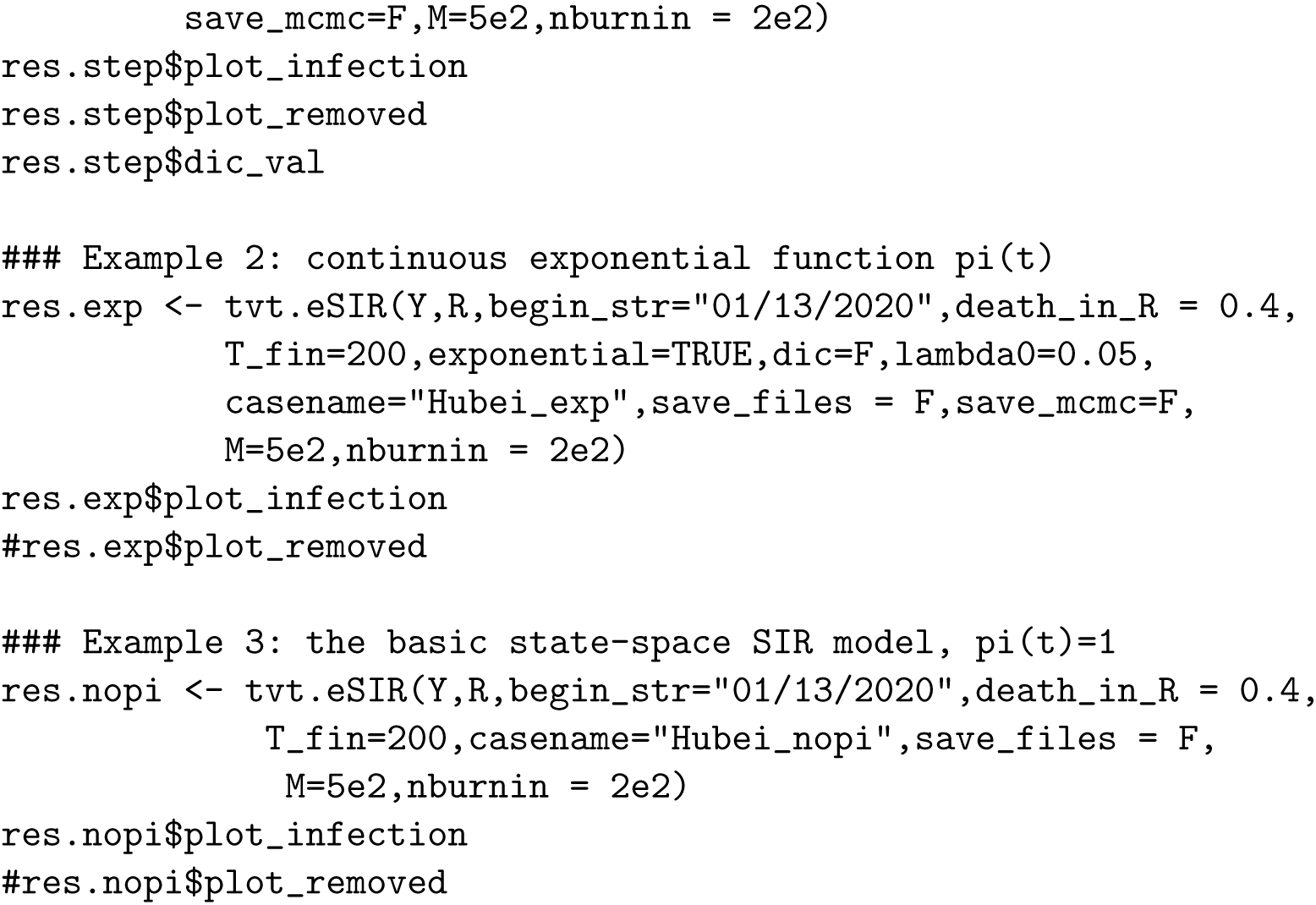

In the above R coding scripts, only very short MCMC chains are specified for the consideration of running time. In practice, it is recommended to set `M=5e5` and `nburnin=2e5` to achieve desirable burn-ins and yield stable MCMC draws. We tried the step function given by Panel C in Figure 3 with `pi0=c(1,0.9,0.5,0.1)`, an exponential function given by Panel B in Figure 4 with rate `lambda0=0.05`, both of which were compared with the basic model with *π*(*t*) = 1. The results of the three modifier functions obtained from the `tvt.eSIR` function are summarized in Figures 9-11. In these forecasting plots of the infected and removed compartments (Panel A and Panel C), the black dots left to the blue vertical line denote the observed proportions of the infected and removed compartments on the last date of available observations or before. That is, the blue vertical marks time *t*_0_ as defined in Section 3. The green and purple vertical lines denote the first and second turning points, respectively. The salmon color area denotes the 95% credible interval of the predicted proportions 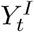 and 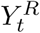 after *t*_0_, respectively, while the cyan color area represents either the 95% credible intervals of the prevalence 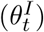 and or that of the probability of removal 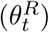 prior to time *t*_0_. The gray and red curves are the posterior mean and median curves. The black curve in the plot (Panel C) is the proportion of deaths estimated from a pre-specified ratio `death_in_R`. The middle Panel B displays important dynamic features of the infection via a spaghetti plot, in which 20 randomly selected MCMC draws of the first-order derivative of the posterior prevalence of infection, namely 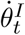. The black curve is the posterior mean of the derivative, and the vertical lines mark times of turning points corresponding respectively to those shown in Panel A and Panel C. Moreover, the 95% credible intervals of these turning points are also highlighted by semi-transparent rectangles in Panel B. As shown in these figures, the transmission rate modifier *π*(*t*) played an important roles in shortening the key turning points of the epidemic, and its effect on both estimation and prediction of the COVID-19 infection dynamics has been clearly demonstrated.

**Figure 9:**
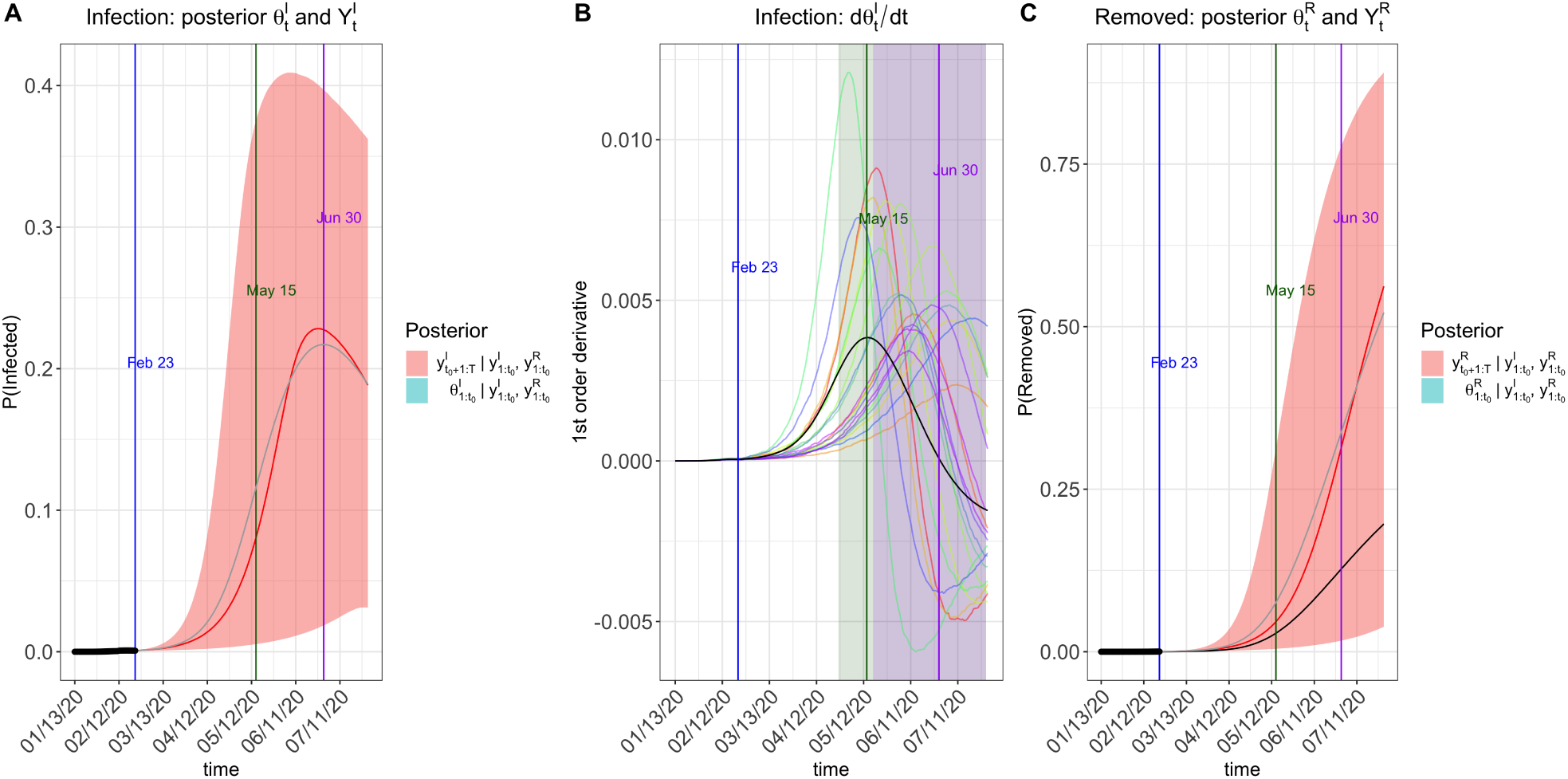
Prediction plots of 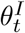 and 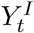 (Panel A), 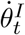 (Panel B), 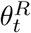 and 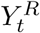 (Panel C) for Hubei without intervention, *π*(*t*) = 1.

**Figure 10:**
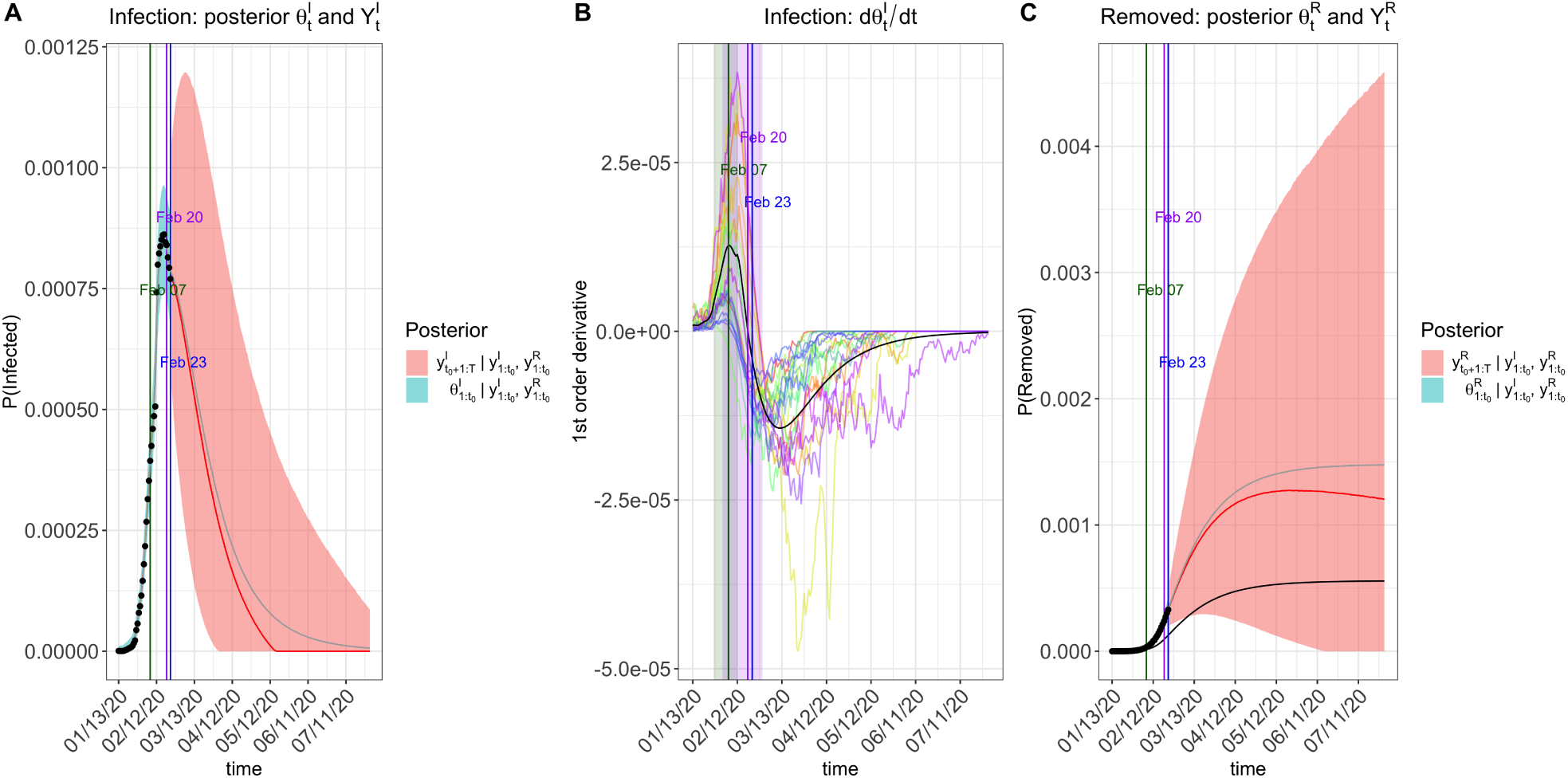
Prediction plots of 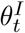 and 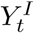 (Panel A), 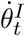 (Panel B), 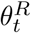 and 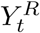 (Panel C) for Hubei with an exponential transmission rate modifier *π*(*t*) = exp)−0.05*t*).

**Figure 11:**
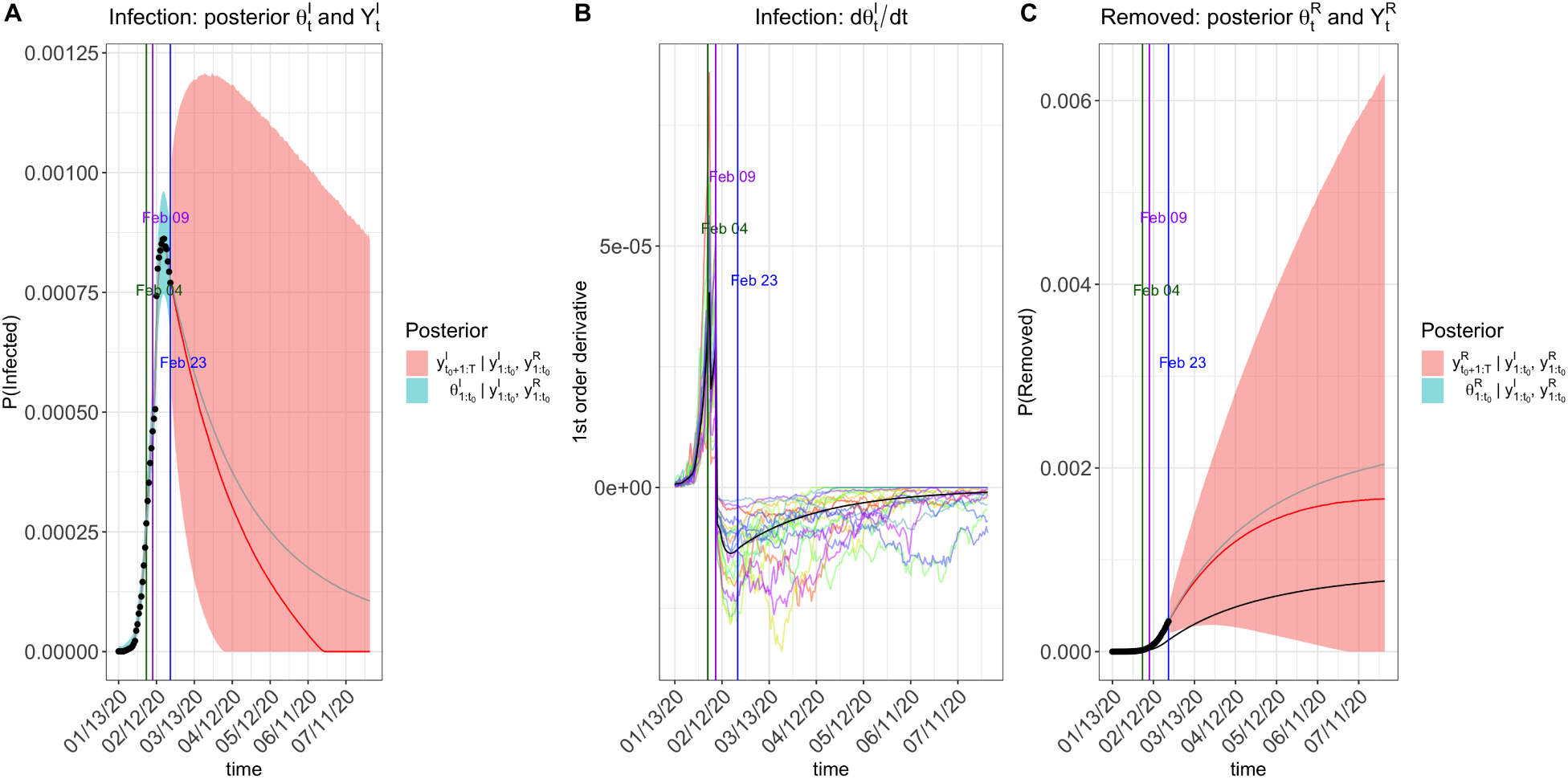
Prediction plots of 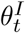 and 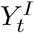 (Panel A), 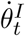 (Panel B), 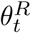 and 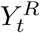 (Panel C) for Hubei with a step-function transmission rate modifier specified by ***π***_0_ = (1, 0.9, 0.5, 0.1) at change points [Jan 23, Feb 4, Feb 8].

Next, we analyzed the data from the rest of the Chinese population (i.e. the provinces outside Hubei) starting on Jan 23. We set `begin_str=“01/23/2020”in tvt.eSIR`. To address the delayed starting time, we included two change points for the step function *π*(*t*) at [Feb 4, Feb 8] with ***π***_0_ = (0.8, 0.1). The exponential function remained the same. It is noted that the spread of

COVID-19 outside Hubei has been so far much less severe. Possible reasons for such low proportions of infection and deaths include (i) discontinuing the traffic connections between Hubei and the provinces, (ii) more timely caution and preventative measures taken, and (iii) a comparatively large population that dilutes the exposed group. When Panel A in Figure 12 is zoomed in, some of the observed proportions (black dots) are deviated from the posterior mean or median of the fitted prevalence albeit they all fall in the 95% credible intervals, as shown by Panel A in Figures 13 and 14. Since the latent process follows the SIR differential equations, there may be a lack of fit for the SIR model to accommodate a very large and complex population of 1.3 billion people, in which most of the subjects are not at risk. The proposed models should work much better for individual provinces, but we did not perform such analyses.

**Figure 12:**
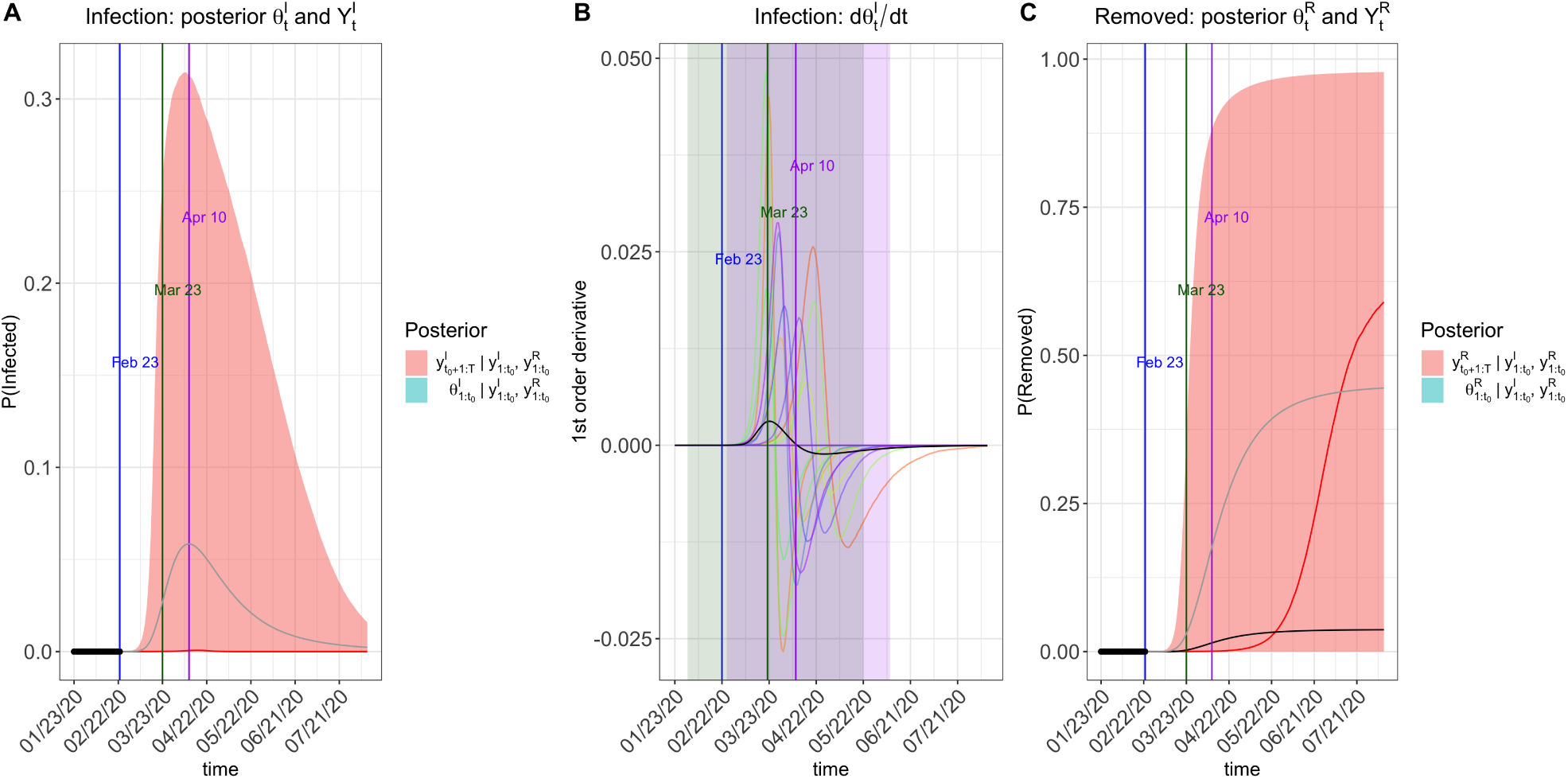
Prediction plots of 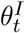 and 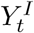 (Panel A), 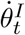 (Panel B), 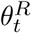 and 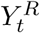 (Panel C) for the other provinces outside Hubei with *π*(*t*) = 1.

**Figure 13:**
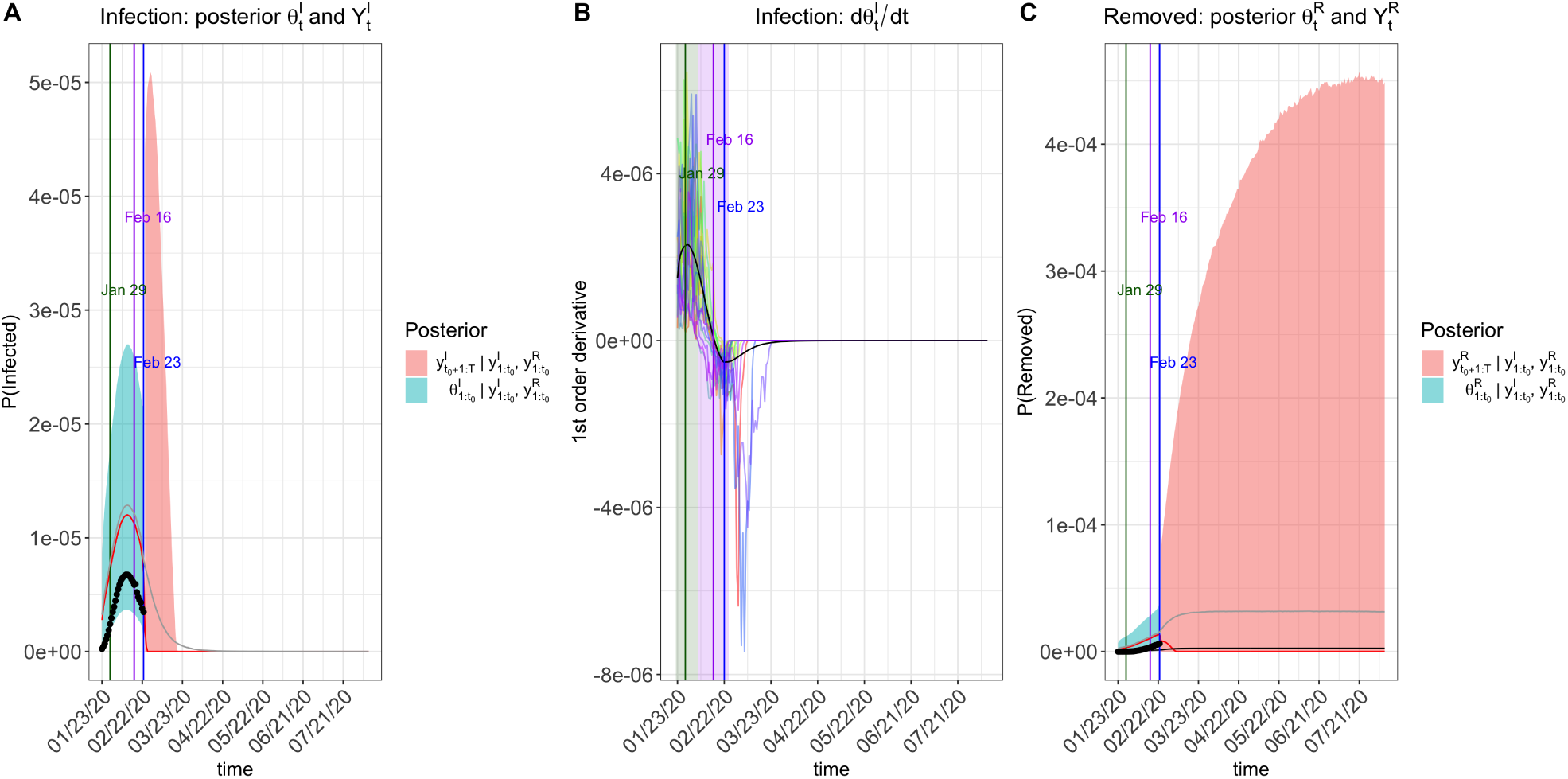
Prediction plots of 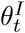 and 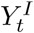 (Panel A), 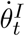 (Panel B), 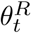 and 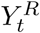 (Panel C) for the other provinces outside Hubei with with an exponential transmission rate modifier *π*(*t*) = exp(−0.05*t*).

**Figure 14:**
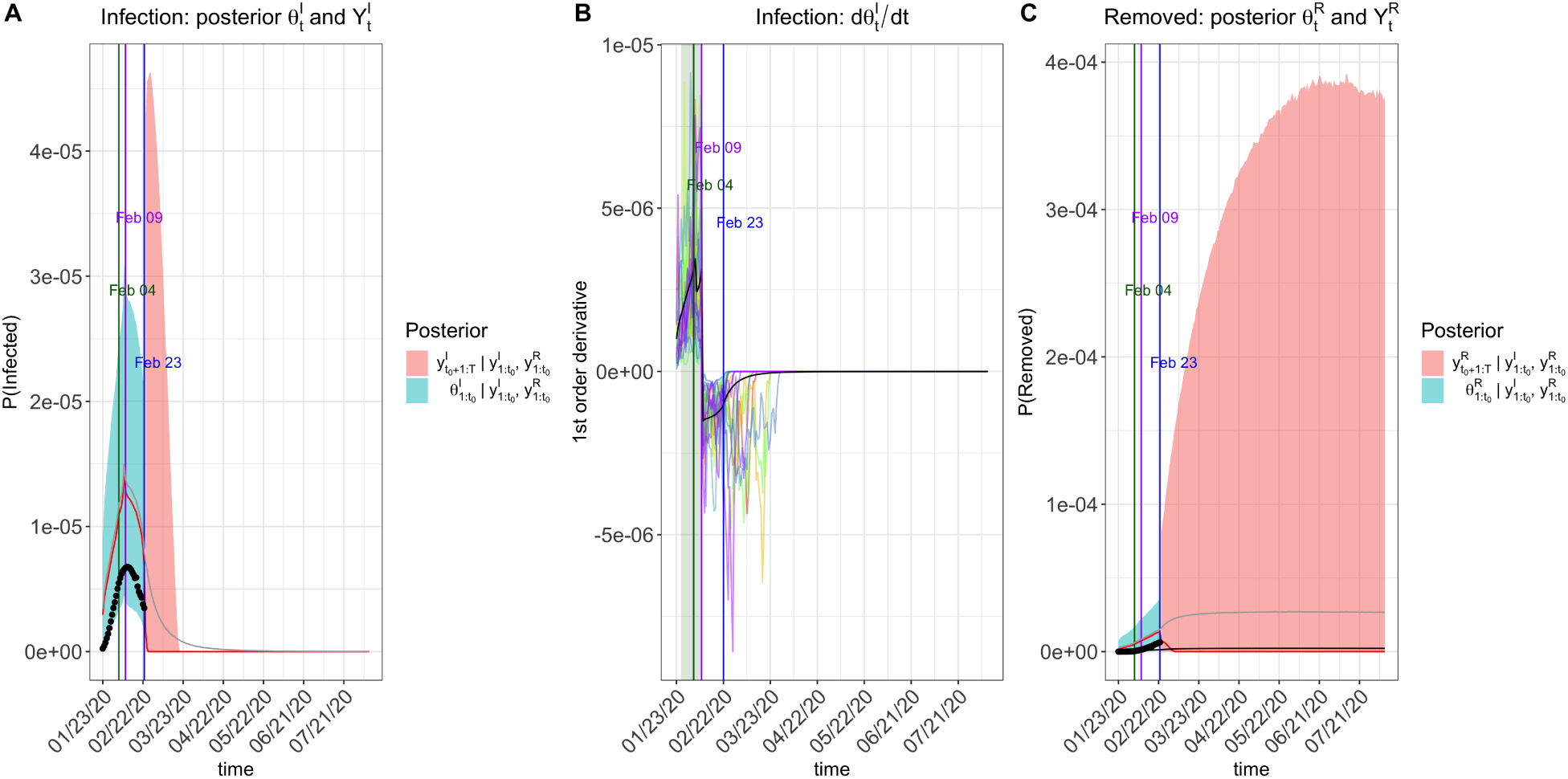
Prediction plots of 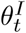 and 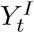 (Panel A), 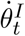 (Panel B), 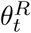 and 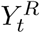 (Panel C) for the other provinces outside Hubei with a step-function transmission rate modifier specified by ***π***_0_ = (1, 0.9, 0.5, 0.1) at change points [Jan 23, Feb 4, Feb 8].

The other epidemiological model with an added quarantine compartment as an absorbing state was fitted via our R function `qh.eSIR` in the package `eSIR`. The arguments used in `qh.eSIR()` are almost identical to those in `tvt.eSIR()`. Note that if the quarantine rate function is set at constant 0, this model will be reduced to a basic epidemiological SIR model.

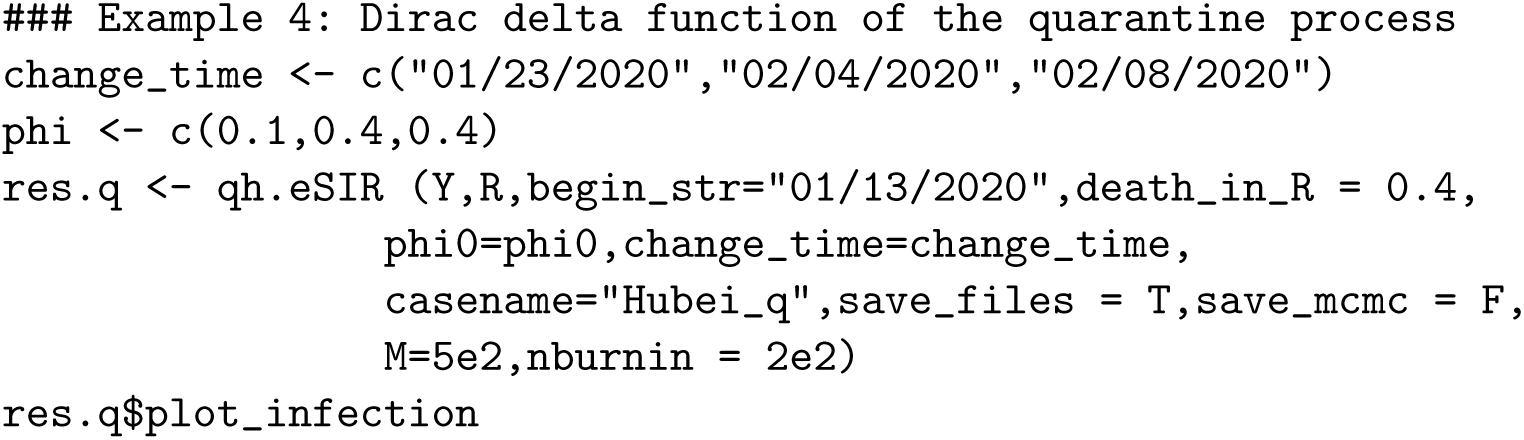
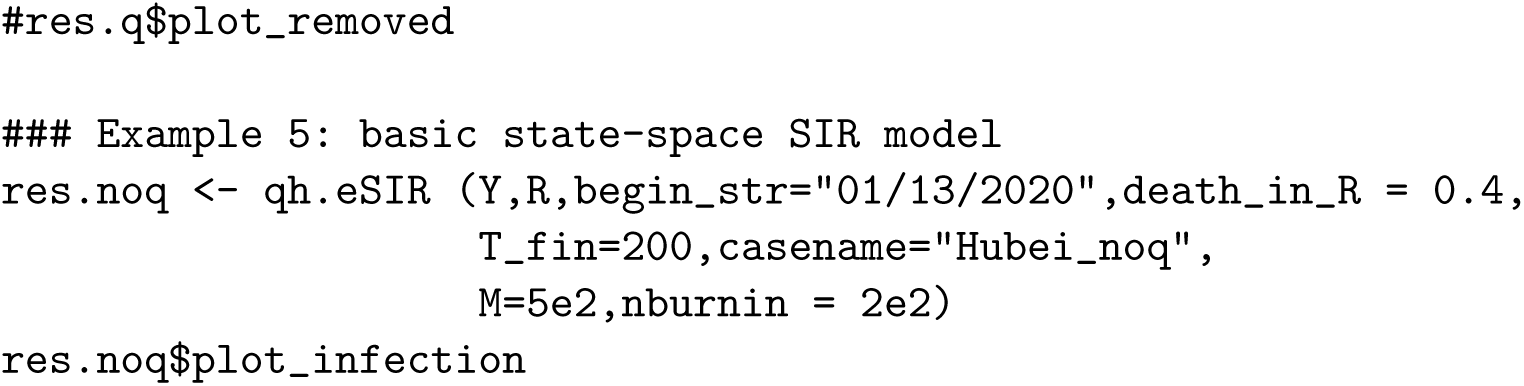

We applied the R function `qh.eSIR` in analyses of the data within and outside Hubei. Results are summarized in Figures 15-16. Our analyses once again clearly indicated that stringent quarantine protocols can largely reduce the spread of COVID-19 both within Hubei and outside Hubei. Yet, it is known that too strict quarantine can cause backfire; people may lose their trust and patience in their committed system, and consequently may try to reduce compliance or even avoid quarantine. We also present the posterior mean probability of staying quarantine compartment in Figure 17 within Hubei and outside Hubei. Note that Jan 23 was not set as a change point for the case of outside Hubei, leading only to two jumps. It is evident that by Feb 8, more than 90% of the Chinese population have taken in-home isolation or as such, reflective to a very strict quarantine protocol enforced in the entire country.

**Figure 15:**
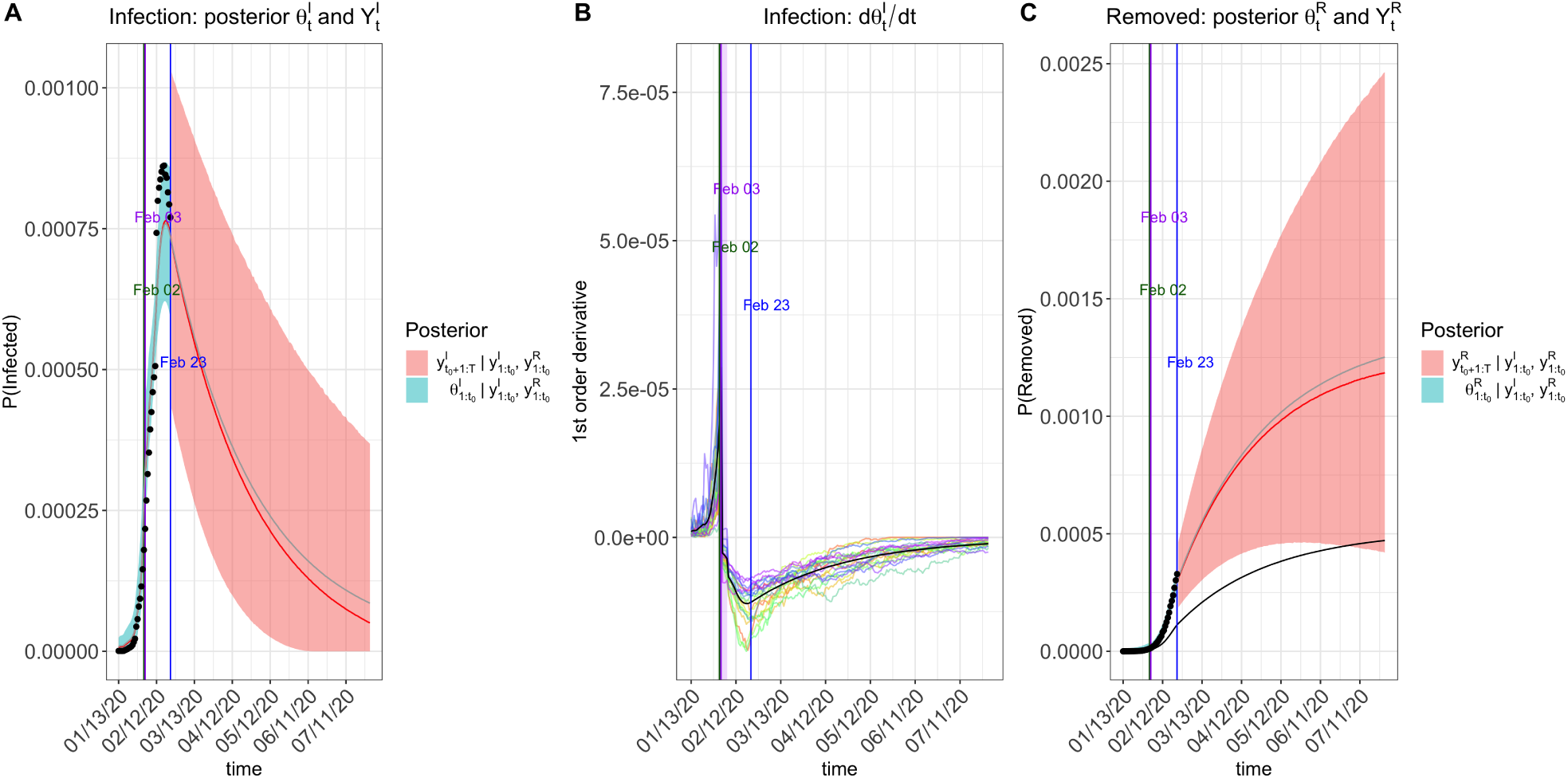
Prediction plots of 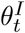 and 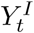 (Panel A), 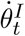 (Panel B), 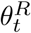 and 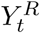 (Panel C) for Hube with time-varying quarantine specified by ***ϕ***_0_ = [0.1, 0.9, 0, 5] at change points [Jan 23, Feb 4, Feb 8].

**Figure 16:**
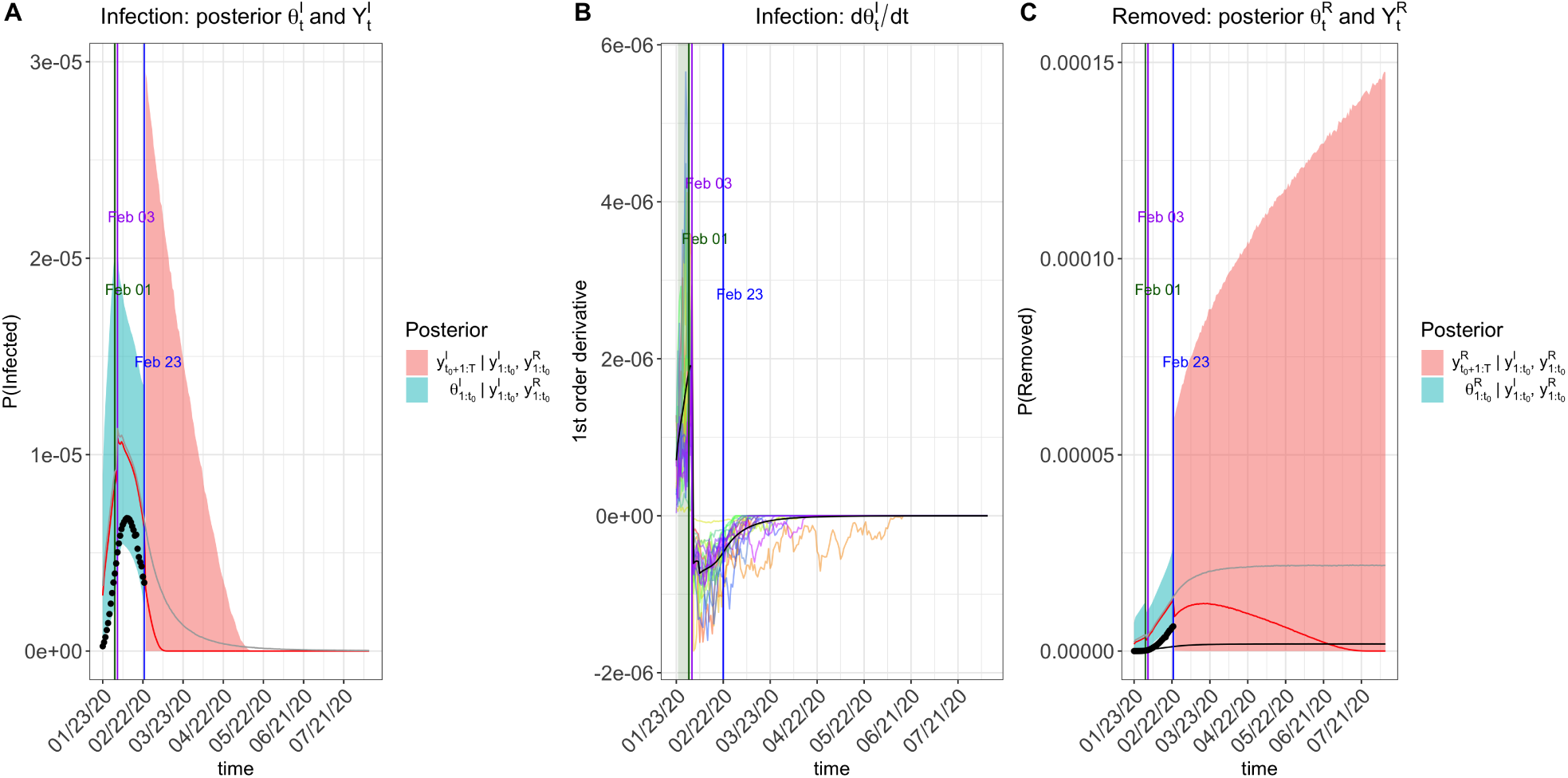
Prediction plots of 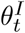 and 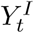 (Panel A), 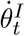 (Panel B), 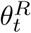 and 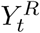 (Panel C) for other provinces outside Hubei with time-varying quarantine specified by ***ϕ***_0_ = [0.9, 0, 5] at change points [Feb 4, Feb 8].

**Figure 17:**
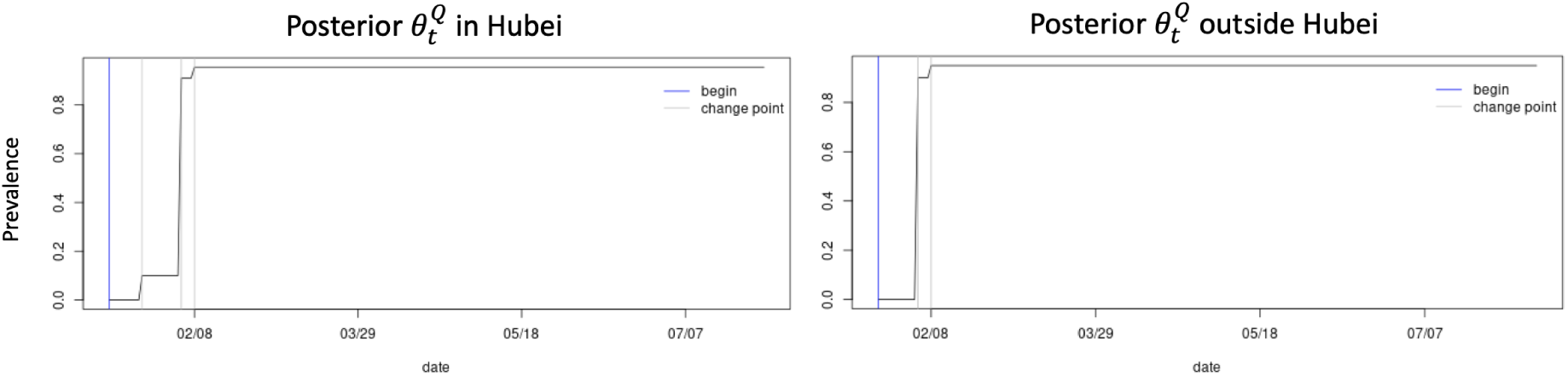
The posterior mean probability of staying in quarantine compartment within and outside Hubei.

As described in Subsection 4.1, we corrected the under-reported proportion of infections in Hubei province prior to Feb 12, when a big jump occurred on one day. We repeated the same analyses using the calibrated data of infections, and the corresponding results are shown in Figures 18-21. It is interesting to see that the abrupt rise in the infection proportion on Feb 12 disappeared in all Panel A in these new analyses, and the observed data (i.e. the black dots) align better with the credible intervals of all the latent processes. We also notice that after the correction, the estimated reproduction numbers *R*_0_ became smaller. The reproduction numbers estimated from different models for within and outside Hubei, with and without the data calibration, together with their 95% credible intervals are summarized in Table 1.

**Table 1:** The posterior mean and credible intervals of the reproduction number *R*_0_ obtained from different quarantine models and datasets.

| Model | Within Hubei |  |  |  | Outside Hubei |  |
| --- | --- | --- | --- | --- | --- | --- |
|  | No Data Calibration |  | Data Calibration |  | Mean | 95%CI |
|  | Mean | 95%CI | Mean | 95%CI |  |  |
| No quarantine | 3.00 | [1.91, 4.50] | 2.96 | [1.82, 4.49] | 2.58 | [1.52, 4.26] |
| Exponential | 6.30 | [2.80, 10.78] | 4.88 | [2.35, 8.42] | 3.36 | [1.96, 5.26] |
| Step-function | 5.61 | [2.89, 8.95] | 4.31 | [2.34, 6.90] | 3.02 | [1.74, 4.89] |
| Quar. Compartment | 6.00 | [2.35, 13.24] | 5.04 | [2.30, 9.36] | 3.58 | [1.85, 6.10] |

**Figure 18:**
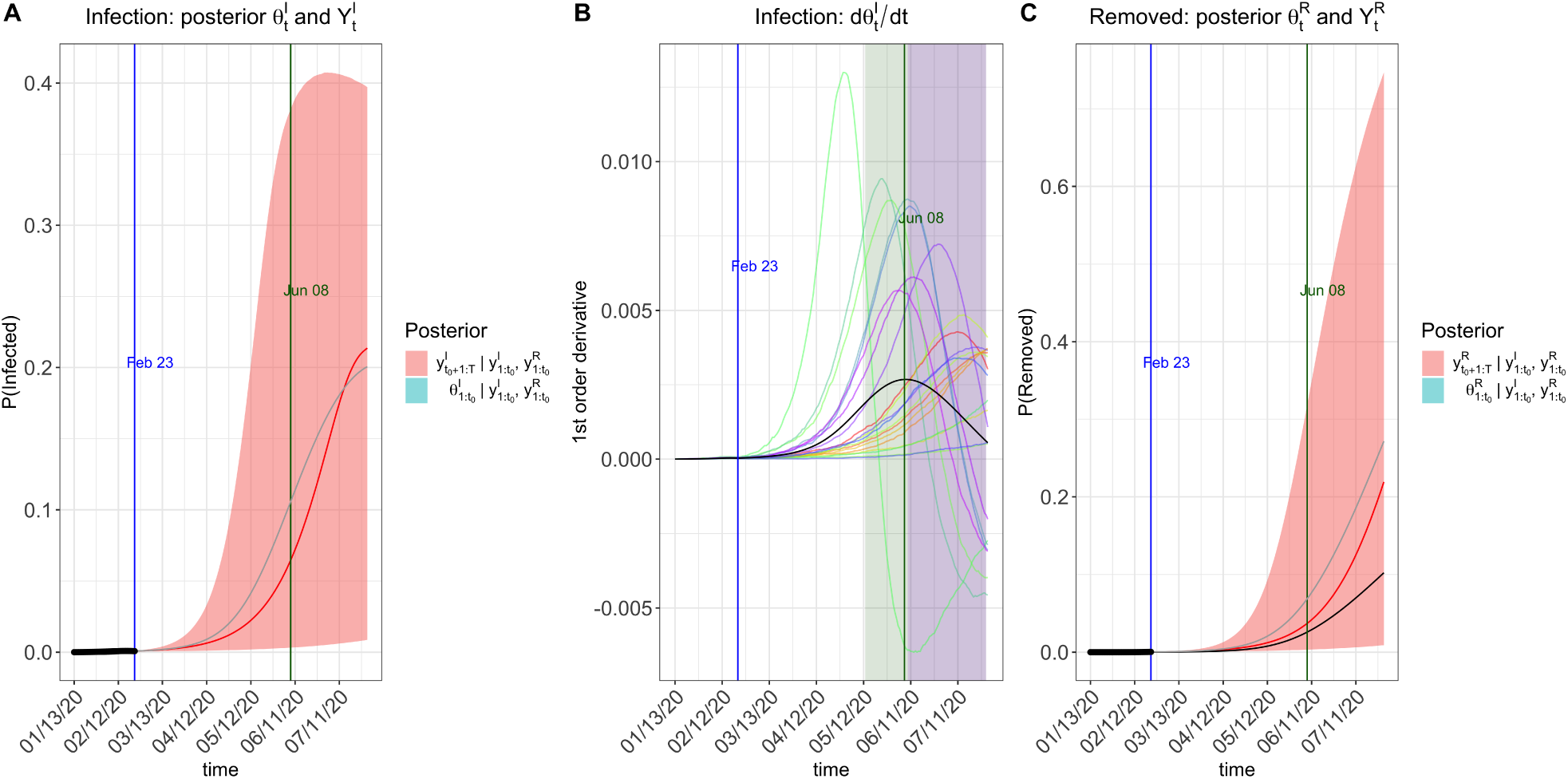
Prediction plots of 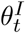 and 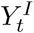 (Panel A), 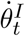 (Panel B), 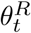 and 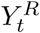 (Panel C) for Hubei province with *π*(*t*) = 1 after calibration. Note that the second turning point is beyond the time-axis limit in the plots.

**Figure 19:**
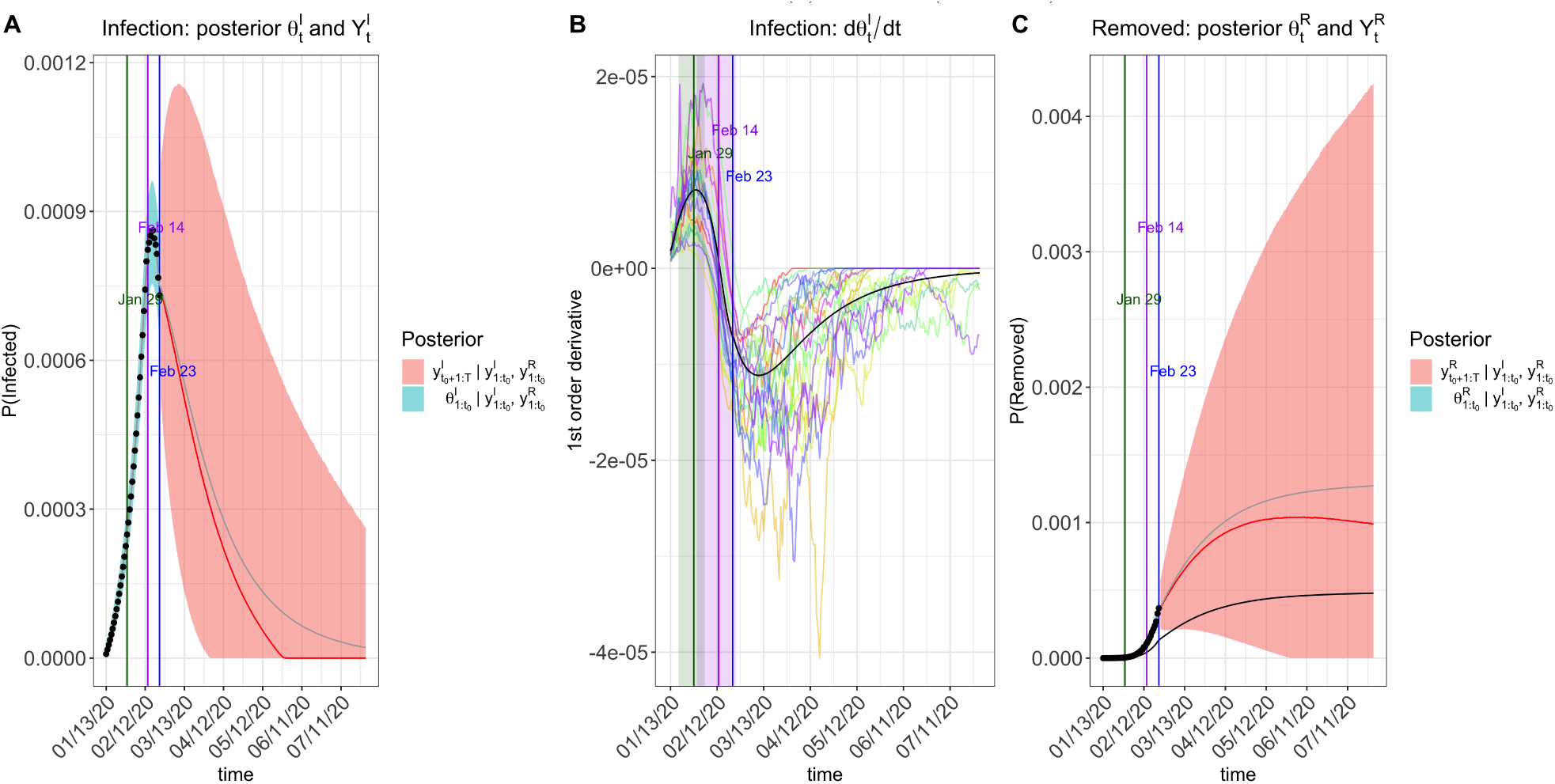
Prediction plots of 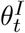 and 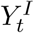 (Panel A), 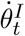 (Panel B), 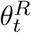and 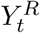, (Panel C) for Hubei with an exponential transmission rate modifier *π*(*t*) = exp(−0.05*t*) after data calibration.

**Figure 20:**
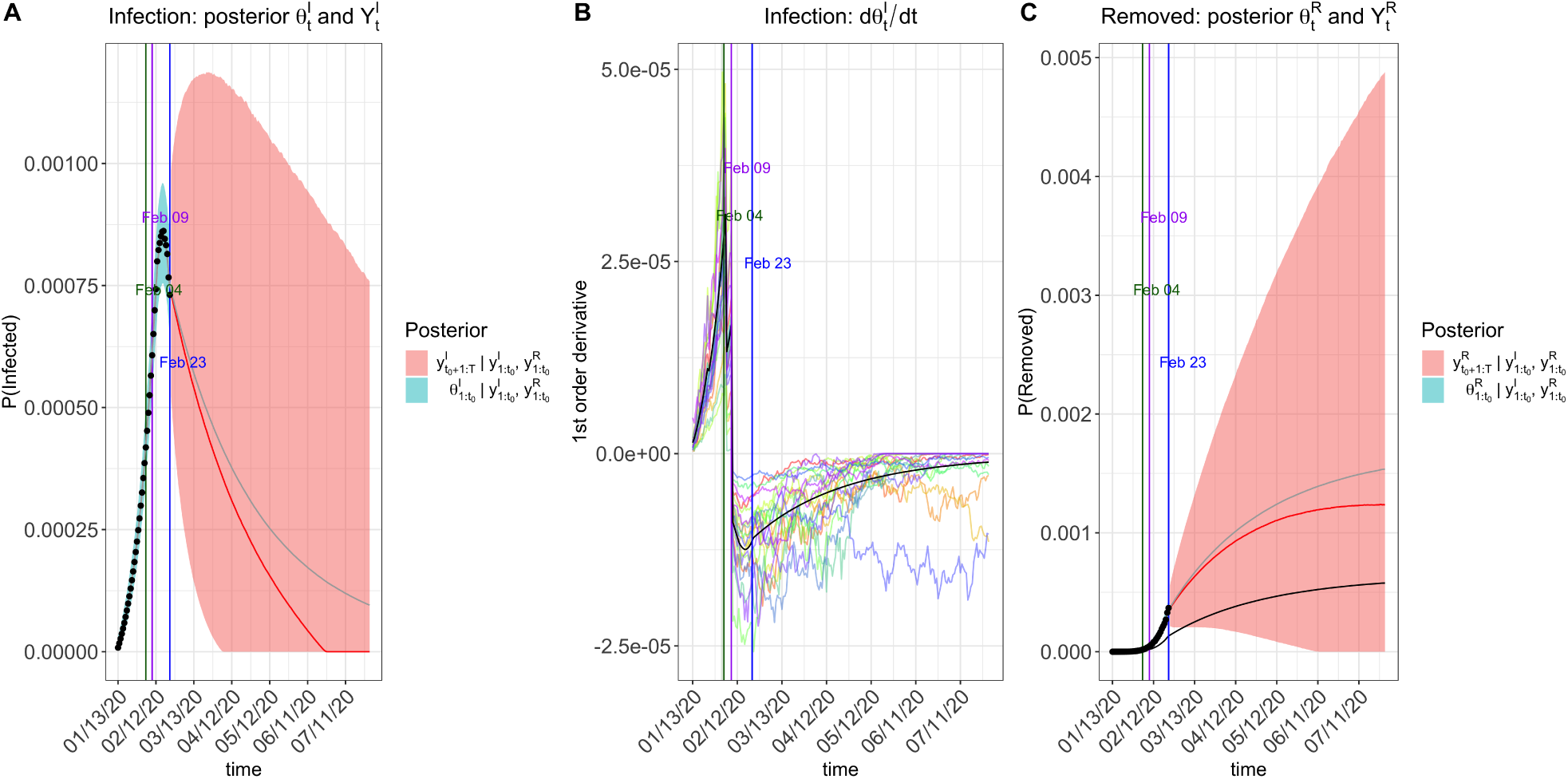
Prediction plots of 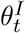 and 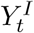 (Panel A), 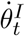 (Panel B), 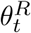 and 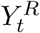 (Panel C) for Hubei with a step-function transmission rate modifier specified by ***π***_0_ = (1, 0.9, 0.5, 0.1) at change points [Jan 23, Feb 4, Feb 8] after data calibration.

**Figure 21:**
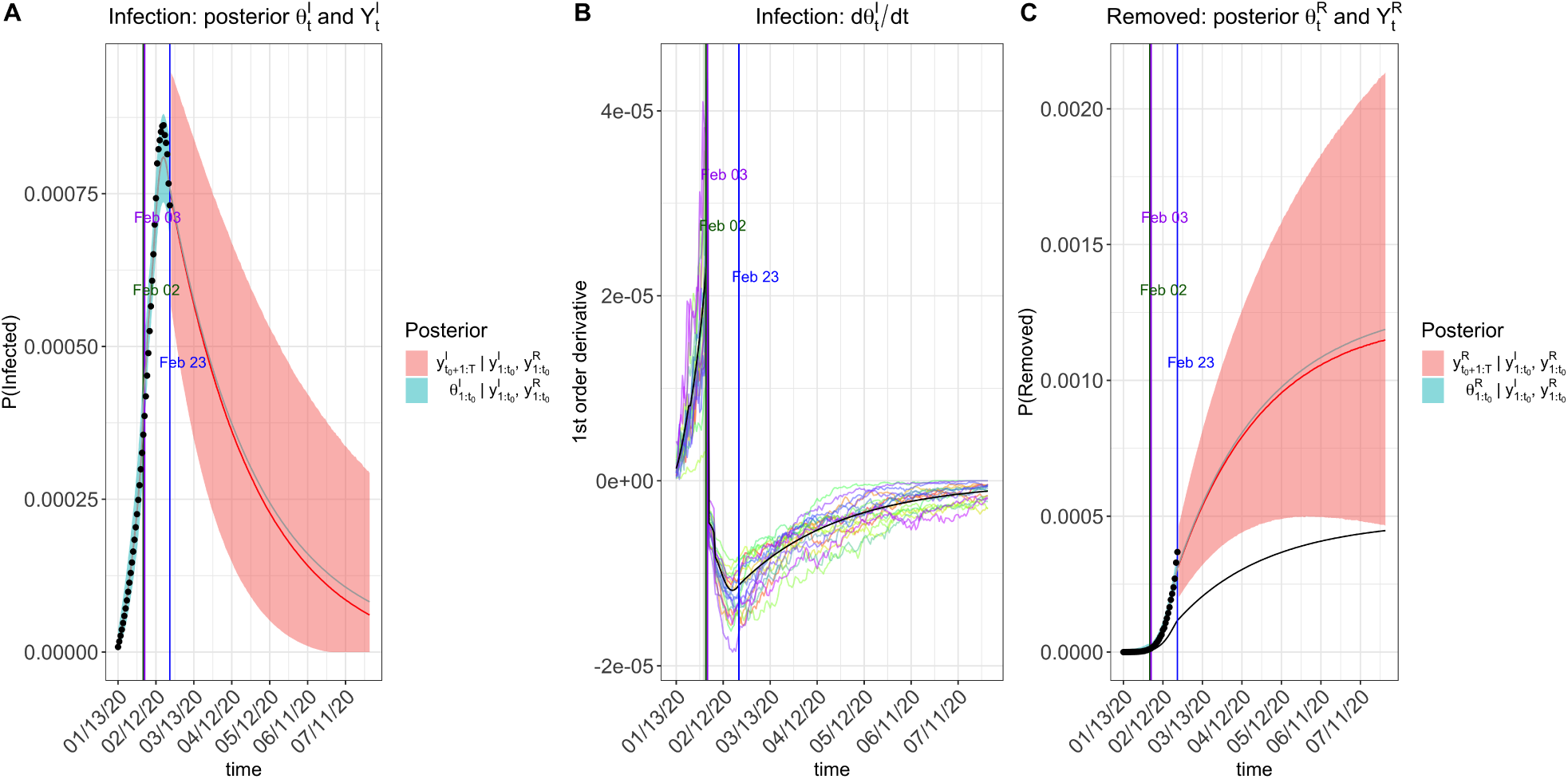
Prediction plots of 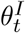 and 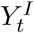 (Panel A), 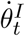 (Panel B), 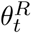 and 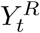 (Panel C) for Hubei with time-varying quarantine specified by ***ϕ***_0_ = [0.1, 0.9, 0, 5] at change points [Jan 23, Feb 4, Feb 8] after data calibration.

It is worth pointing out that the estimates of the reproduction numbers obtained from the epidemiological models with time-varying transmission or quarantine rates appear larger than those obtained from the basic model with no quarantine. This is no surprising as our new models explicitly incorporate interventions, so that the estimated *R*_0_ is an adjusted number with the influence of interventions be removed. In contrast, the basic model with no use of the quarantine modifier implicitly integrates the effect of interventions into the transmission rate *β*, and consequently the estimated *R*_0_ is reduced due to the contribution from interventions. Our analyses suggest that reproduction numbers *R*_0_ of COVID-19 without public health interventions would be around 4-5 within Hubei and around 3-3.5 outside Hubei with relatively big credible intervals. These findings are in agreement with findings from [17]. As pointed out above, the size of credible interval may be reduced with more accessible data of COVID-19, which permit users to specify smaller variances in the prior distributions given in section 3.1.

## 5 Concluding Remarks

We develop an epidemiological forecast model with an R software package to assess effects of interventions on the COVID-19 epidemic within Hubei and outside Hubei in China. Since our proposed model utilizes the strength of the SIR’s dynamic system to capture the primary mechanism of the COVID-19 infectious disease, we are able to predict future episodes of the disease spread patterns over a window of 200 days from the last date of data availability. Some turning points of interest are obtained from these forecasting curves as part of the deliverable information, including the predicted time when daily proportion of infected cases becomes smaller than the previous ones and the predicted time when daily proportion of removed cases (i.e. both recovered and dead) becomes larger than that of infected cases, as well as the time when the epidemic ends. Our informatics toolbox provides quantification of uncertainty on the prediction, rather than only point prediction values, which are valuable to see the best versus the worst. The key novel contribution is the incorporation of time-varying quarantine protocols to expand the basic epidemiological model to accommodate changing transmission rates over time in the population. The toolbox can be used by practitioners who have better knowledge of quarantine and better quality data to perform their own analyses. Practitioners can use the toolbox to evaluate different types of quarantine strategies in practice. All summary statistics obtained from the toolbox are of great importance for public health workers and government policy makers to take proper actions on stop spreading of COVID-19.

We chose the MCMC algorithm to implement the statistical estimation and prediction because of the consideration on the prediction uncertainty. Given the considerable complexity in the COVID-19 virus spread dynamics and potentially inaccurate information of quarantine measures as well as likely under-reported proportions of infected and recovered cases and deaths, it is of critical importance to quantify and report uncertainty in the forecast. Note that the publicly reported data of recovery and death of COVID-19 are mostly collected from hospitals where accessibility to such information is warrant. In contrast, it is very difficulty, if not impossible, to collect the data of infected individuals with light symptoms who had in-home isolation and recovered, in spite of serious efforts made by the government for a door-to-door inspection to identify suspected cases.

This toolbox is indeed so general that it may be applicable to analyze and evaluate the COVID-19 epidemic in other countries, as well as the future outbreak of other types of infectious diseases. As noted in the paper, our proposed method does need prior data of similar infectious disease to set up initial conditions of the infection dynamics. For this, we analyzed the complete SARS data from Hong Kong given some similarity of COVID-19 to SARS. From this perspective, what we learned from this COVID-19 epidemic in this paper is extremely valuable to form initial conditions in the analysis of any future outbreak of similar infectious disease. In addition, understanding forms and strengths of quarantines for the controlling of disease spread is an inevitable path to making effective preventive policies, which is the key analytic capacity that our toolbox offers.

The proposed epidemiological models can be further extended to accommodate more data reported by the China CDC, which are worth future exploration. Two types of data that may be used in the future extension are the daily number of suspected cases and the daily number of hospitalized cases. We did not use such data due to the concern of data accuracy. For example, the number of suspected cases is largely dependent on the diagnostic protocols, which have been revised a few times since the outbreak of the disease, and the sensitivity of the RNA test. Given such concerns, our strategy in the proposed model was to only use necessary data for analysis, and over the course of improved data quality in the near future, our toolbox may be extended to enjoy greater statistical power and more accurate predictions.

## Data Availability

We used the publicly available data from the China CDC, which can be accessed at DXY.cn.

## Supplementary Material

Software website: https://github.com/lilywang1988/eSIR

## A Runge-Kutta Approximation

### A.1 Approximation in the Basic SIR model

The forth order Runge-Kutta(RK4) method gives an approximate of *f* (***θ***_*t*−_, *β, γ*) in equation (4) as follows:

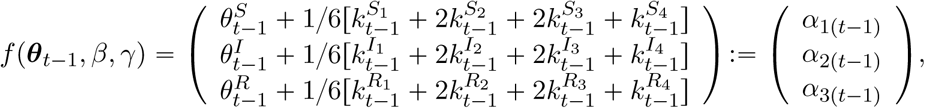

where

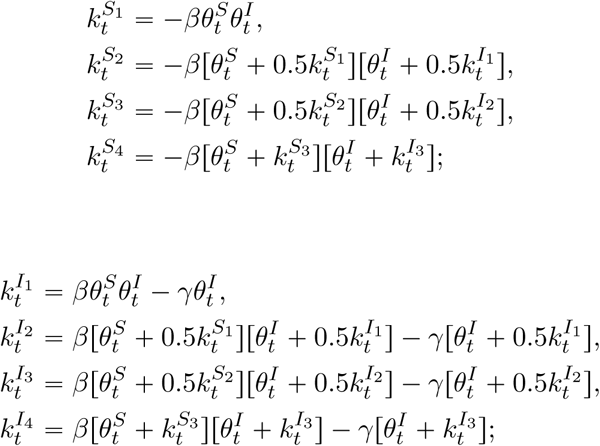

and

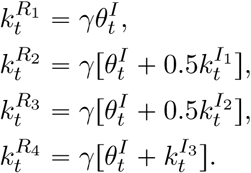

### A.2 Approximation in the extended SIR model with quarantine compartment

Using the RK4 approximation, *f* (***θ***_*t*−_, *β, γ*) in the extended SIR model (6) with a quarantine compartment can be approximated following the two iterative steps:

1. Solve the *f* (***θ***_*t*−_, *β, γ*) in Appendix A without considering the quarantine with *f*(·)

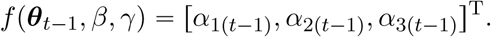

2. Due to the quarantine, we deduct the susceptible by 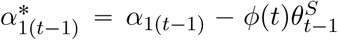, and let 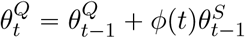 with 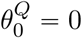.

Let 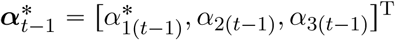, and it is easy to show that the sum 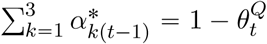. Thus we can regenerate the next day’s ***θ***_*t*_ following a Dirichlet distribution adjusted by the prevalence of the quarantine compartment 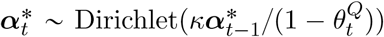. The estimated prevalence values become 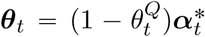. We follow above two steps and finish the complete prevalence processes. Note that the deduction of susceptible compartments might cause 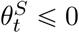, we will bound such prevalence value to be consistently 0, which is equivalent to terminating transmission among susceptible subjects.

### B Moment properties of Beta and Dirichlet distributions

For the sake of being self-contained, we list the moments of both Beta and Dirichlet distributions. The mean and variance of Beta distribution Beta(*α, β*) are respectively:

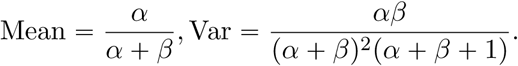

While to Dirchlet distribution Dir(*κ****α***), we have

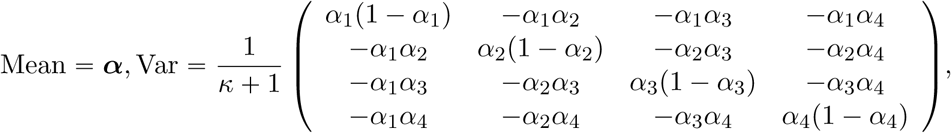

where ***α*** = (*α*_1_, *α*_2_, *α*_3_, *α*_4_)^T^ with *α*_1_ *α*_2_ *α*_3_ *α*_4_ = 1.

### C Under-reporting Calibration

As is mentioned in the Introduction, the issue of under-reporting may cause bias in prediction. In order to adjust the under-reported number of infected cases, we apply the following algorithm to calibrate the number of infections before Feb 12, during which time the Chinese government only relies on the RNA test for diagnosis, which was realized later with low sensitivity leading to many false negatives.

We assume that the cumulative number of infected cases between Jan 13 and Feb 12 when a sudden big jump occurs follows an exponential function,

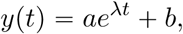

where *t* ∈ {1, 2, …} and *a, b, λ* are parameters to be estimated. Here, *t* = 1 stands for Jan 13 and *t* = 31 stands for Feb 12. Under the condition of *y*(0) = 0, we can easily get that

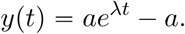

To estimate parameter *λ* and *a*, we want to minimize the least square error of the estimated number *ŷ*(*t*) of infected cases at *t* = 32 (Feb 13), which is one day after the Chinese government changed the diagnosis protocol. It is assumed that the difference between the predicted and observed number of infections on Feb 13 would not be big if the model were established well, although the long term difference might be large due to other interventions. Therefore, the optimization problem we want to solve is,

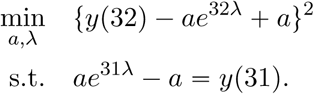

The constraint *ae*^31*λ*^ − *a* = *y*(31) is used to ensure that the cumulative number of infected cases till Feb 12 equals to the observed value *y*(31). The optimization problem can be solved using the method of Lagrange Multipliers. Obtained 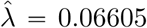, â = 7142.80. The calibrated number of infected cased between Jan 13 and Feb 12 is shown in Figure 8. The proposed calibration method corrected the under-reporting issue, at least partially.

## Package ‘eSIR’

February 29, 2020

**Type** Package

**Title** Extended state-space SIR models

**Version** 0.2.0

**Date** 2020-02-19

**Author** Song Lab (http://www.umich.edu/~songlab/)

**Maintainer** Lili Wang

**Description** An inplementation of extended state-space SIR models developed by Song Lab at UM school of Public Health

**License** CC BY 4.0

**Encoding** UTF-8

**Depends** rjags, scales, ggplot2, chron, gtools, data.table

**RoxygenNote** 6.1.1

## R topics documented

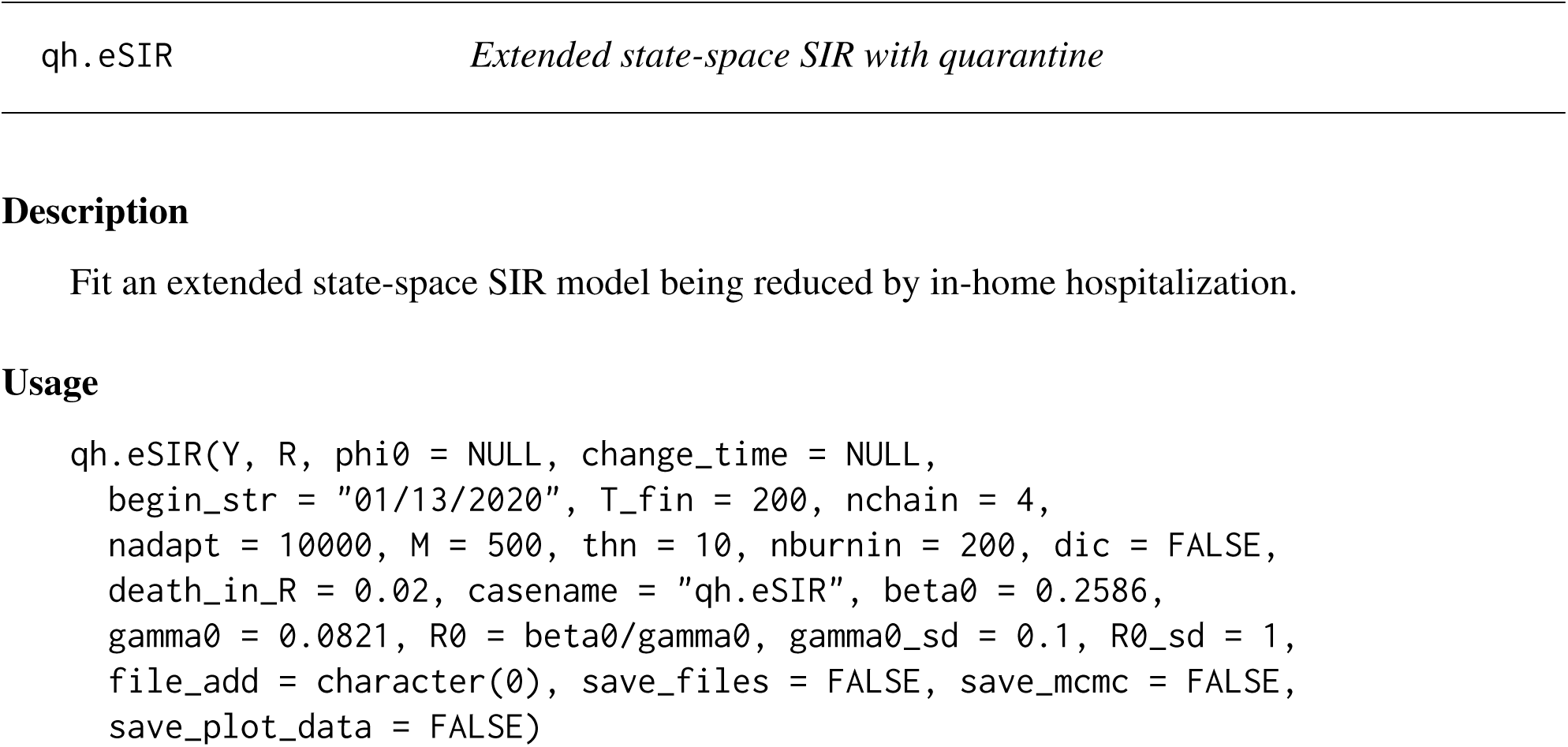

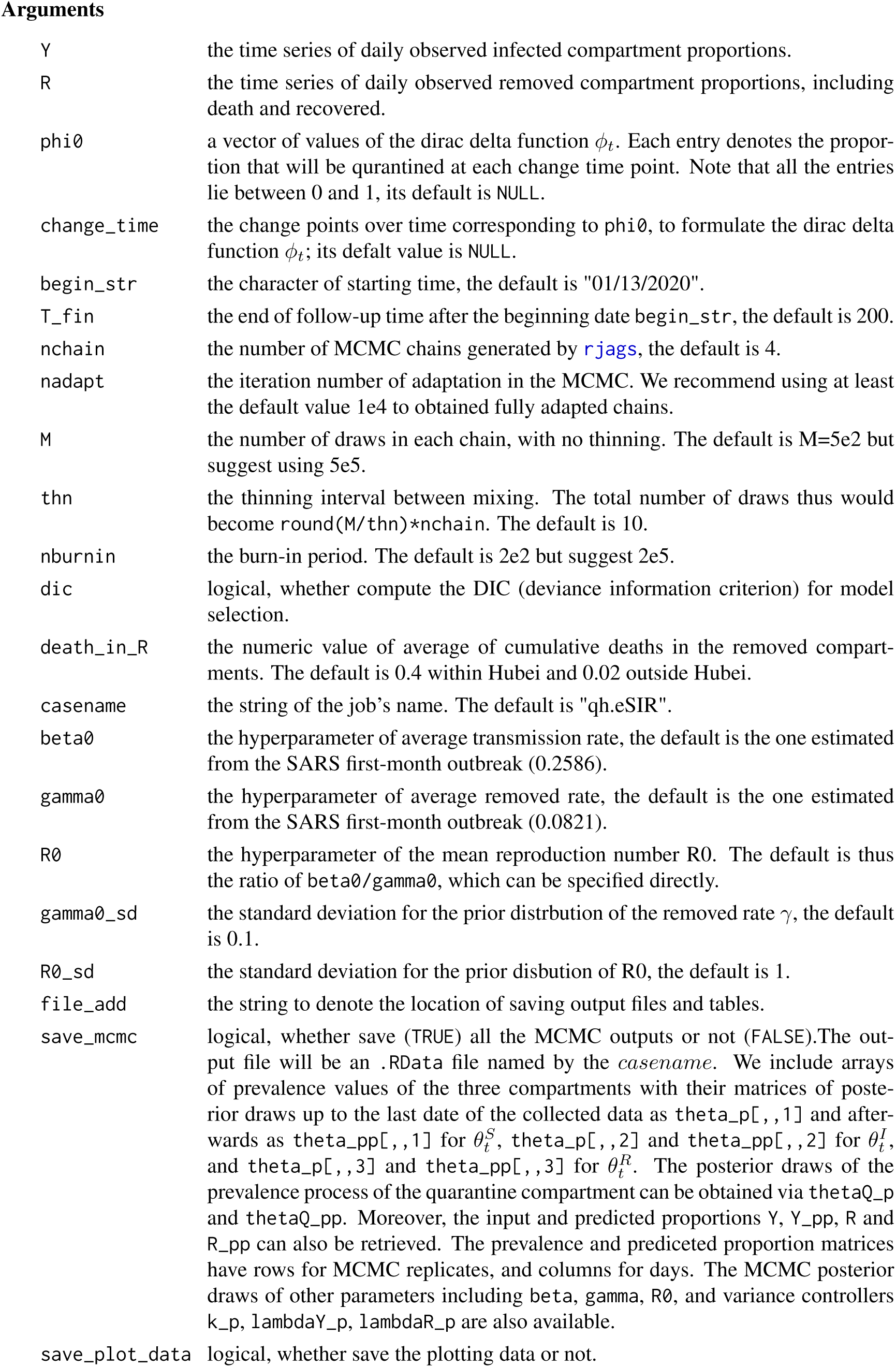

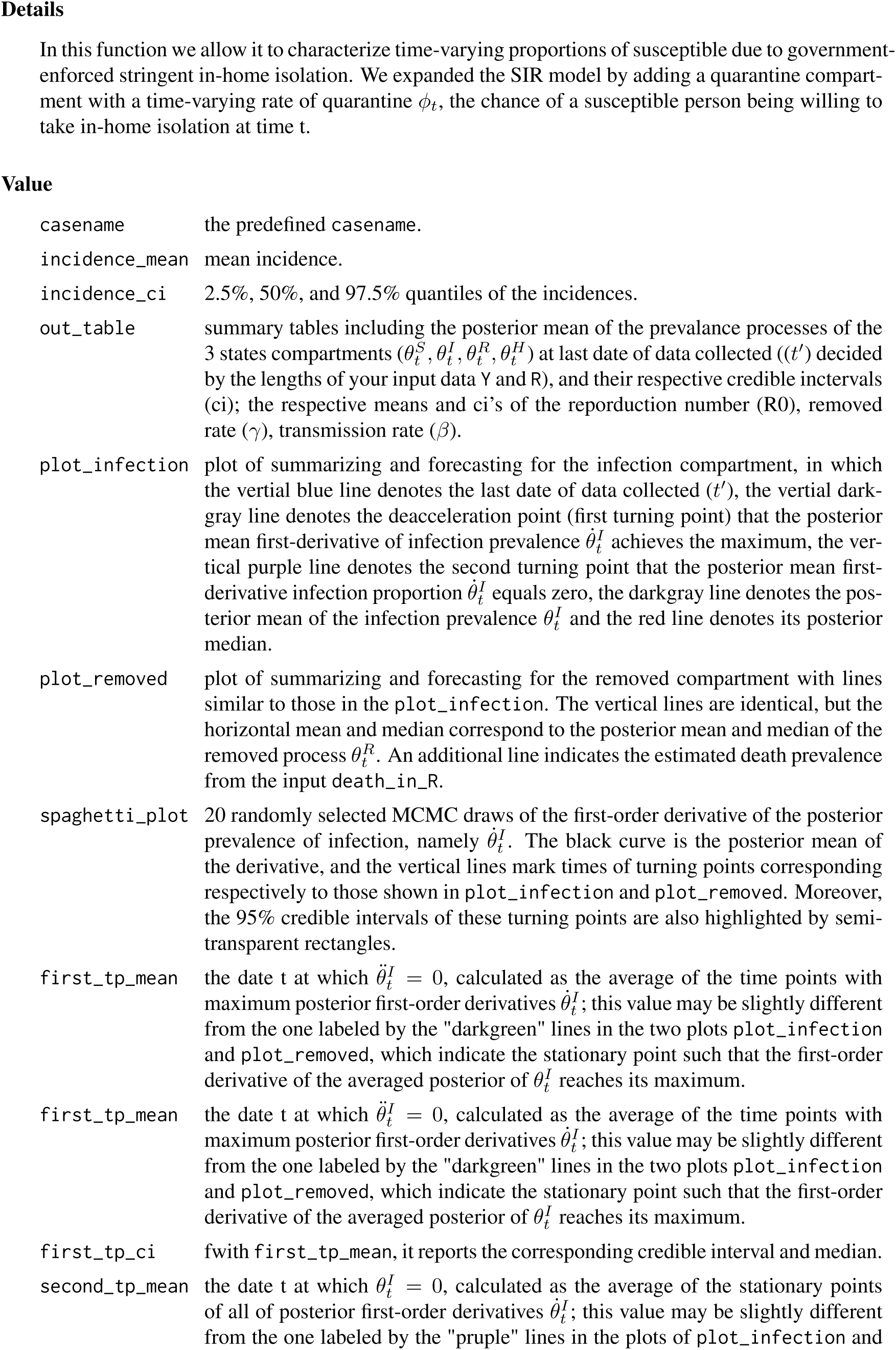

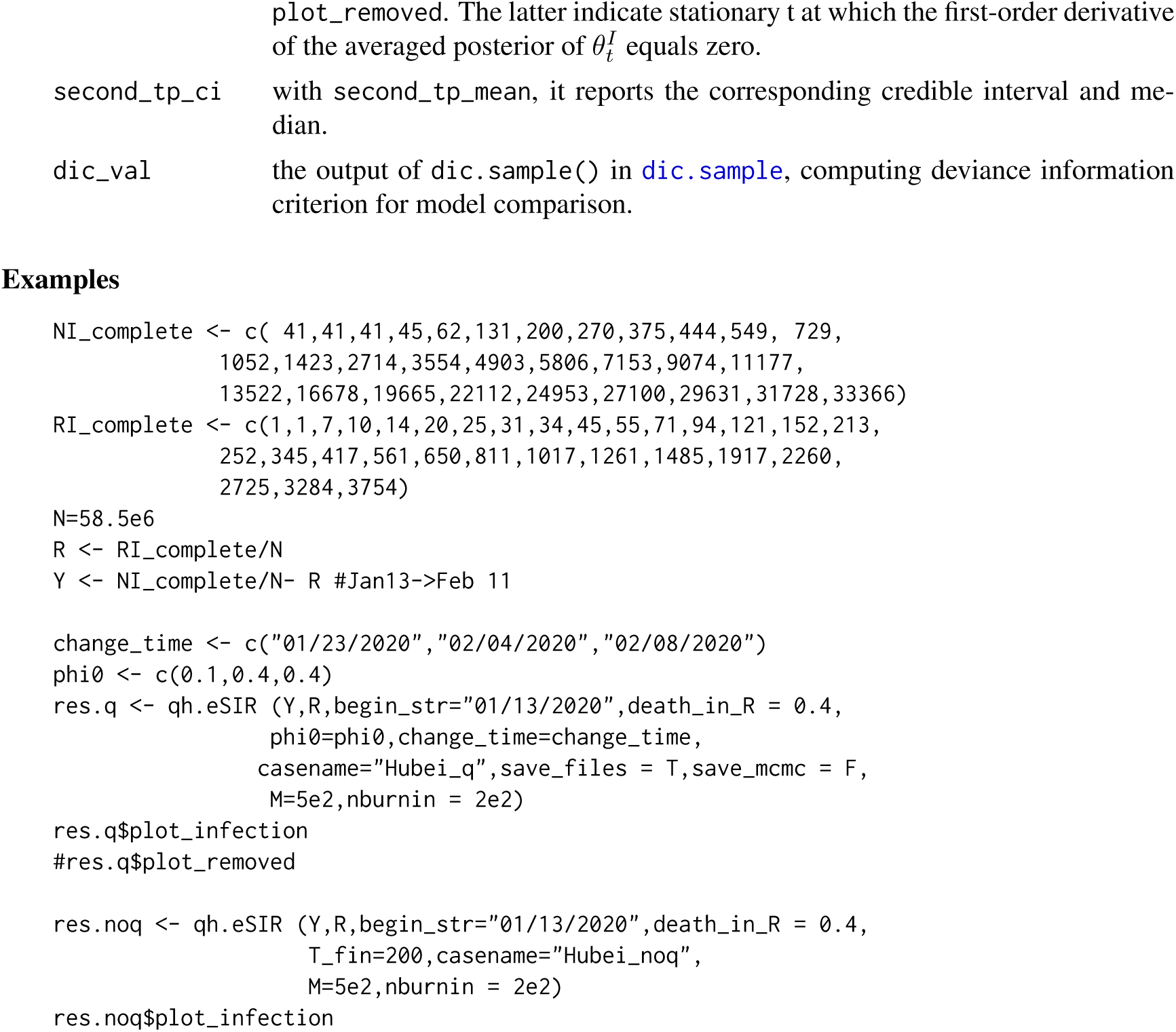

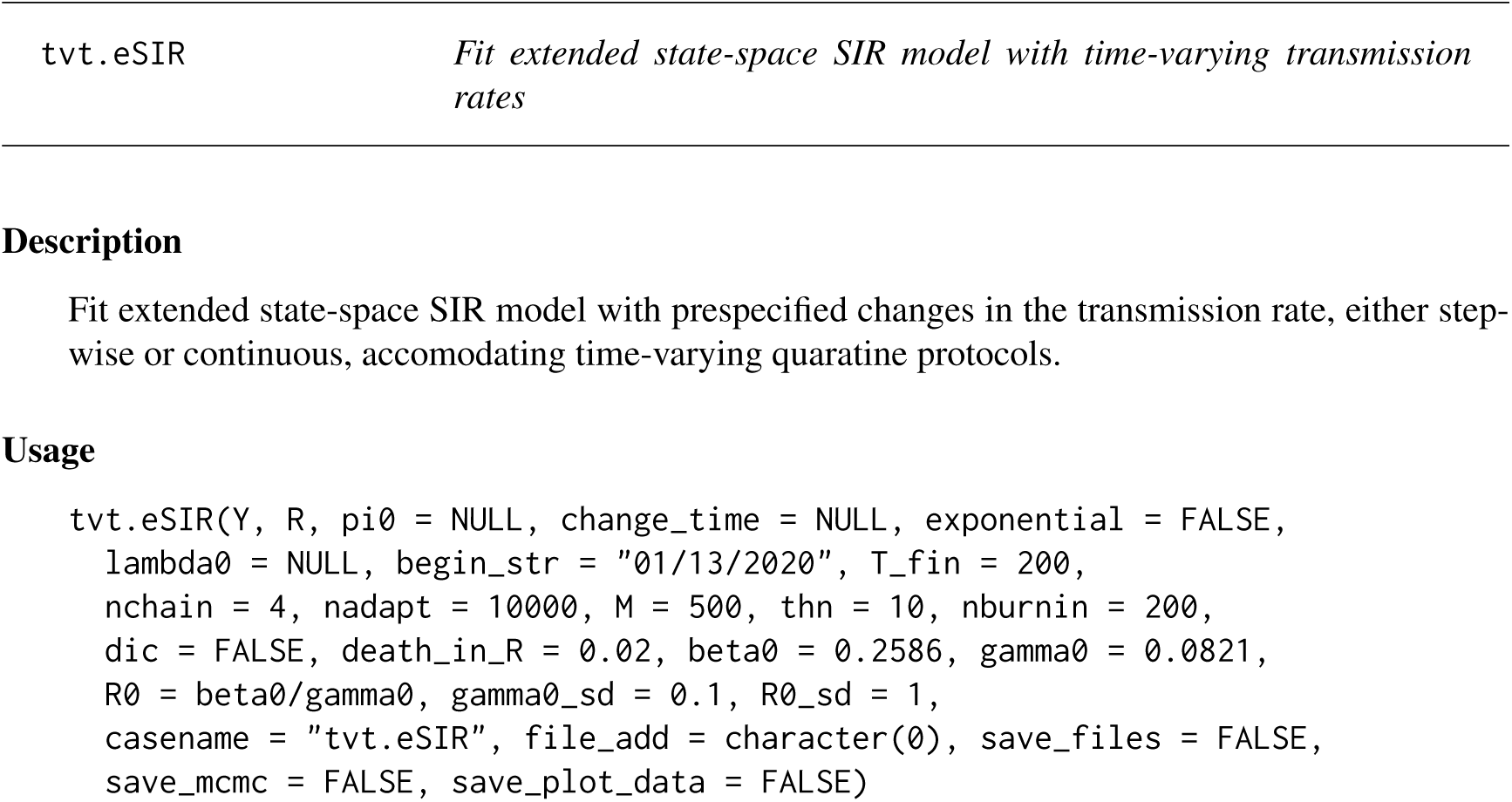

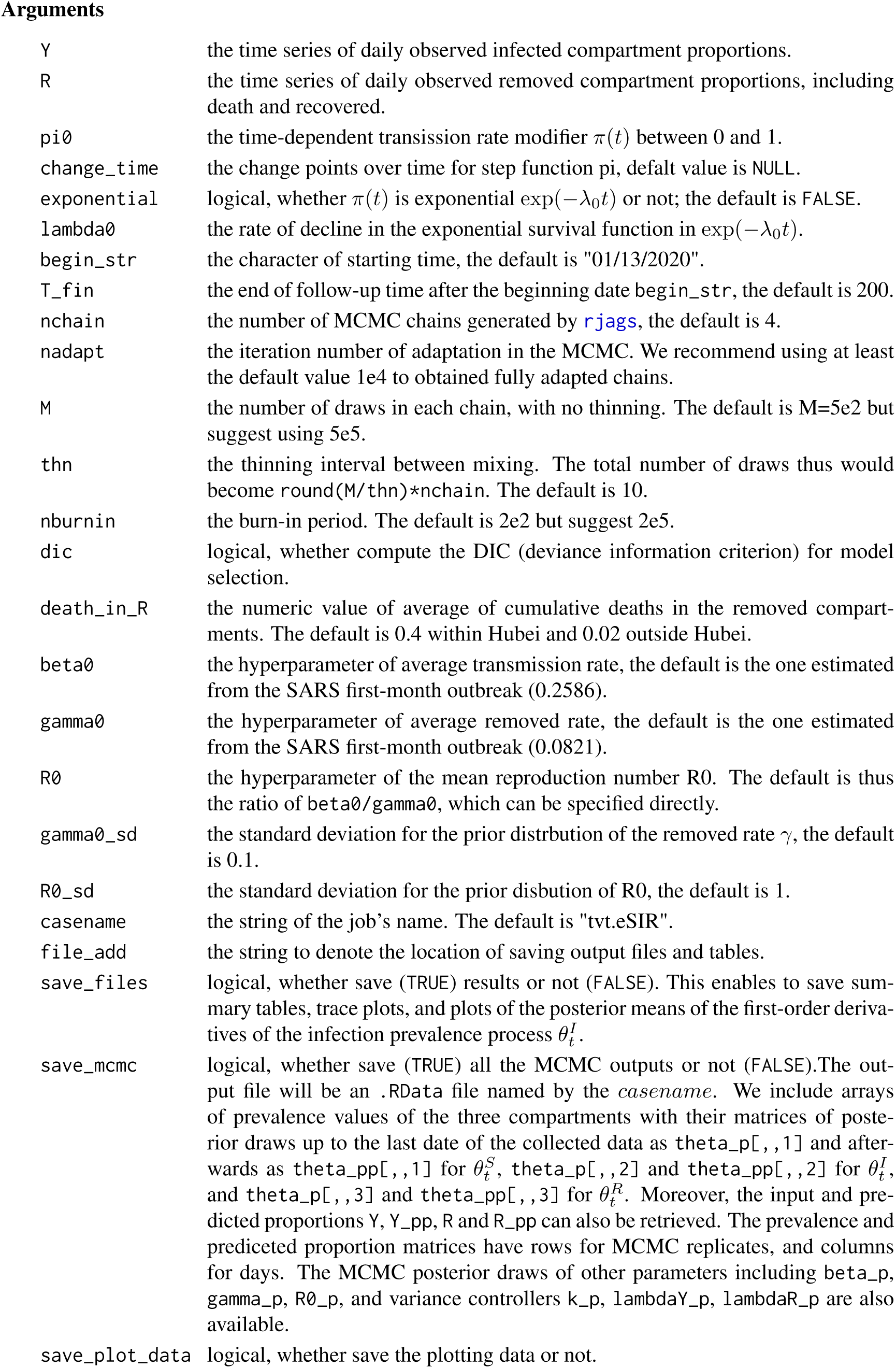

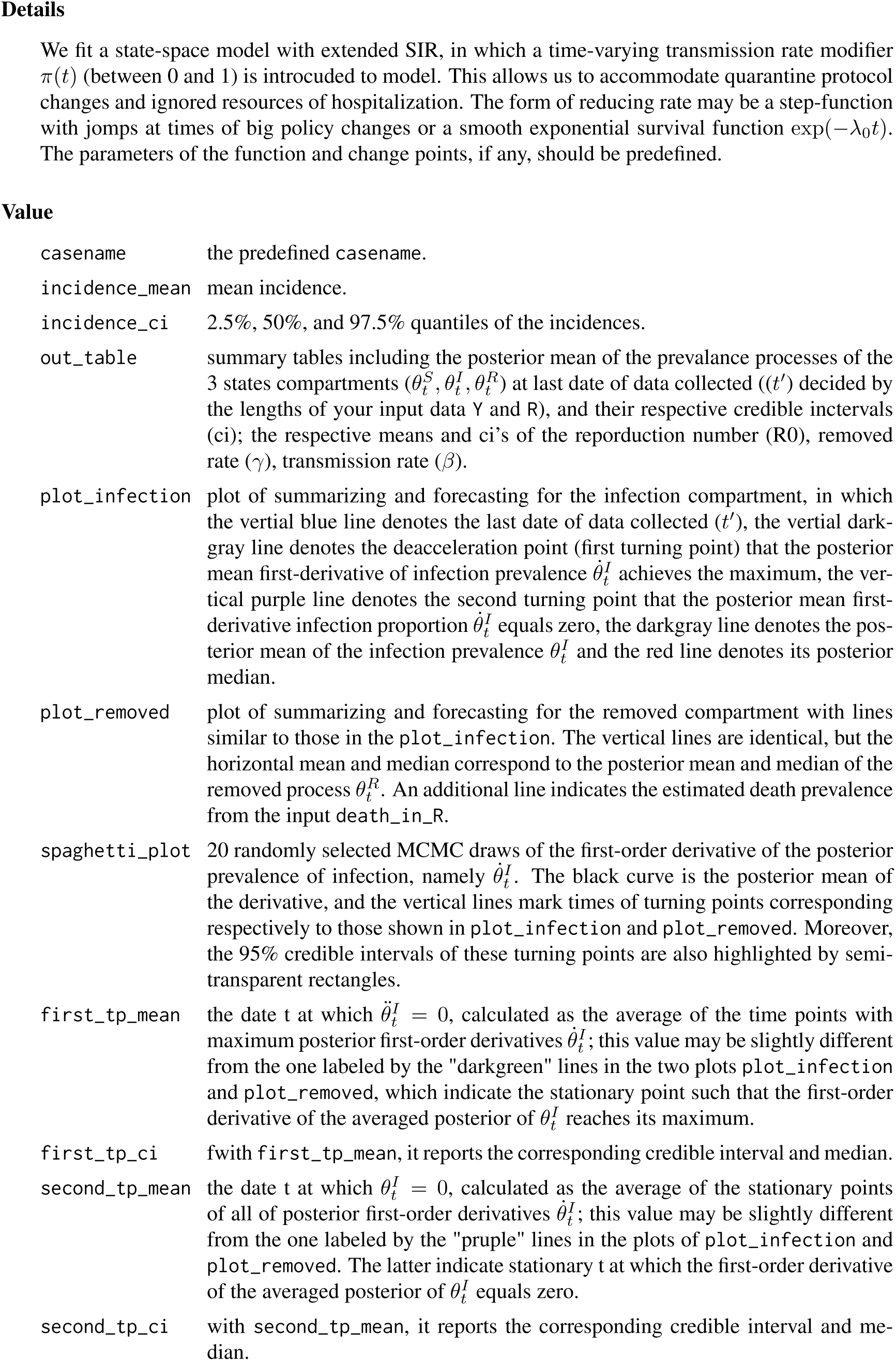

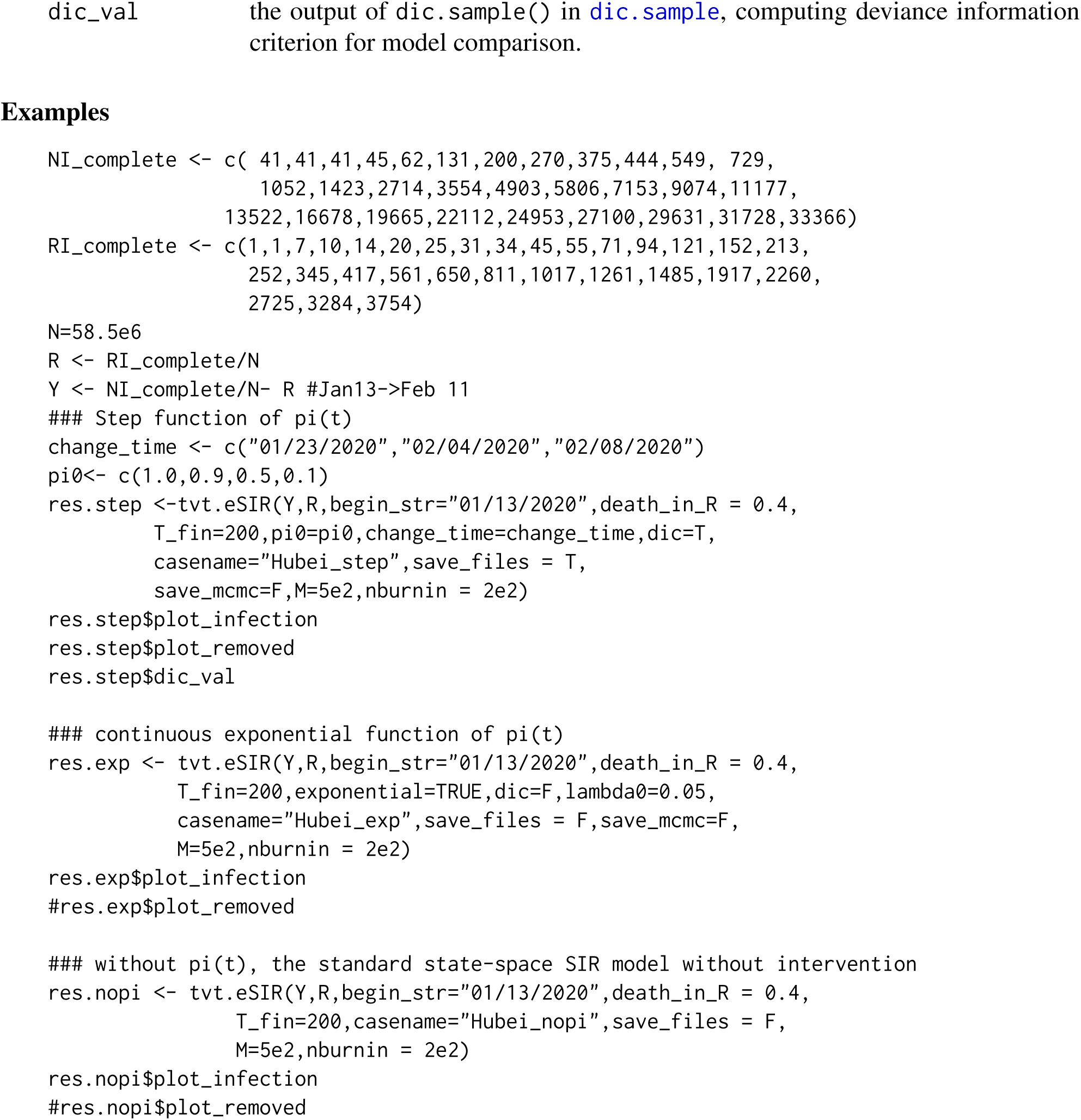

## References

[1] Song Lab at UM. eSIR: Extended state-space SIR models, 2020. R package version 0.1.0.

[2] Bradley P Carlin, Nicholas G Polson, and David S Stoffer. A monte carlo approach to non-normal and nonlinear state-space modeling. Journal of the american Statistical association, 87(418):493–500, 1992.

[3] Huijun Chen, Juanjuan Guo, Chen Wang, Fan Luo, Xuechen Yu, Wei Zhang, Jiafu Li, Dongchi Zhao, Dan Xu, Qing Gong, et al. Clinical characteristics and intrauterine vertical transmission potential of covid-19 infection in nine pregnant women: a retrospective review of medical records. The Lancet, 2020.

[4] David T Dennis, Kenneth L Gage, Norman G Gratz, Jack D Poland, Evgueni Tikhomirov, World Health Organization, et al. Plague manual: epidemiology, distribution, surveillance and control. Technical report, Geneva: World Health Organization, 1999.

[5] Yi Fan, Kai Zhao, Zheng-Li Shi, and Peng Zhou. Bat coronaviruses in china. Viruses, 11(3):210, 2019.

[6] Lisa E Gralinski and Vineet D Menachery. Return of the coronavirus: 2019-ncov. Viruses, 12(2):135, 2020.

[7] Wei-jie Guan, Zheng-yi Ni, Yu Hu, Wen-hua Liang, Chun-quan Ou, Jian-xing He, Lei Liu, Hong Shan, Chun-liang Lei, David SC Hui, et al. Clinical characteristics of 2019 novel coronavirus infection in china. medRxiv, 2020.

[8] Michelle L Holshue, Chas DeBolt, Scott Lindquist, Kathy H Lofy, John Wiesman, Hollianne Bruce, Christopher Spitters, Keith Ericson, Sara Wilkerson, Ahmet Tural, et al. First case of 2019 novel coronavirus in the united states. New England Journal of Medicine, 2020.

[9] Zixin Hu, Qiyang Ge, Li Jin, and Momiao Xiong. Artificial intelligence forecasting of covid-19 in china. arXiv preprint 2002.07112, 2020.

[10] Chaolin Huang, Yeming Wang, Xingwang Li, Lili Ren, Jianping Zhao, Yi Hu, Li Zhang, Guohui Fan, Jiuyang Xu, Xiaoying Gu, et al. Clinical features of patients infected with 2019 novel coronavirus in wuhan, china. The Lancet, 2020.

[11] David S Hui, Esam I Azhar, Tariq A Madani, Francine Ntoumi, Richard Kock, Osman Dar, Giuseppe Ippolito, Timothy D Mchugh, Ziad A Memish, Christian Drosten, et al. The continuing 2019-ncov epidemic threat of novel coronaviruses to global health—the latest 2019 novel coronavirus outbreak in wuhan, china. International Journal of Infectious Diseases, 91:264–266, 2020.

[12] Natsuko Imai, Ilaria Dorigatti, Anne Cori, Steven Riley, and Neil M Ferguson. Estimating the potential total number of novel coronavirus cases in wuhan city, china, 2020.

[13] Bent Jørgensen, Soren Lundbye-Christensen, PX-K Song, and Li Sun. A state space model for multivariate longitudinal count data. Biometrika, 86(1):169–181, 1999.

[14] Bent Jøsrgensen and Peter X-K Song. Stationary state space models for longitudinal data. Canadian Journal of Statistics, 35(4):461–483, 2007.

[15] Sung-mok Jung, Andrei R Akhmetzhanov, Katsuma Hayashi, Natalie M Linton, Yichi Yang, Baoyin Yuan, Tetsuro Kobayashi, Ryo Kinoshita, and Hiroshi Nishiura. Real-time estimation of the risk of death from novel coronavirus (covid-19) infection: Inference using exported cases. Journal of Clinical Medicine, 9(2):523, 2020.

[16] William Ogilvy Kermack and Anderson G McKendrick. A contribution to the mathematical theory of epidemics. Proceedings of the royal society of london. Series A, Containing papers of a mathematical and physical character, 115(772):700–721, 1927.

[17] Jinghua Li, Yijing Wang, Stuart Gilmour, Mengying Wang, Daisuke Yoneoka, Ying Wang, Xinyi You, Jing Gu, Chun Hao, Liping Peng, et al. Estimation of the epidemic properties of the 2019 novel coronavirus: A mathematical modeling study. 2020.

[18] Qiang Li and Wei Feng. Trend and forecasting of the covid-19 outbreak in china. arXiv preprint 2002.05866, 2020.

[19] Qinghe Liu, Zhicheng Liu, Deqiang Li, Zefei Gao, Junkai Zhu, Junyan Yang, and Qiao Wang. Assessing the tendency of 2019-ncov (covid-19) outbreak in china. medRxiv, 2020.

[20] Hayes KH Luk, Xin Li, Joshua Fung, Susanna KP Lau, and Patrick CY Woo. Molecular epidemiology, evolution and phylogeny of sars coronavirus. Infection, Genetics and Evolution, 2019.

[21] Thembinkosi Mkhatshwa and Anna Mummert. Modeling super-spreading events for infectious diseases: case study sars. arXiv preprint 1007.0908, 2010.

[22] World Health Organization et al. Summary of probable sars cases with onset of illness from 1 november 2002 to 31 july 2003. http://www.who.int/csr/sars/coun-try/table20040421/en/index.html, 2003.

[23] Dave Osthus, Kyle S Hickmann, Petruţa C Caragea, Dave Higdon, and Sara Y Del Valle. Forecasting seasonal influenza with a state-space sir model. The annals of applied statistics, 11(1):202, 2017.

[24] Liangrong Peng, Wuyue Yang, Dongyan Zhang, Changjing Zhuge, and Liu Hong. Epidemic analysis of covid-19 in china by dynamical modeling. arXiv preprint 2002.06563, 2020.

[25] Martyn Plummer. rjags: Bayesian Graphical Models using MCMC, 2019. R package version 4-10.

[26] R Core Team. R: A Language and Environment for Statistical Computing. R Foundation for Statistical Computing, Vienna, Austria, 2018.

[27] Jomar F Rabajante. Insights from early mathematical models of 2019-ncov acute respiratory disease (covid-19) dynamics. arXiv preprint 2002.05296, 2020.

[28] Camilla Rothe, Mirjam Schunk, Peter Sothmann, Gisela Bretzel, Guenter Froeschl, Claudia Wallrauch, Thorbjörn Zimmer, Verena Thiel, Christian Janke, Wolfgang Guggemos, et al. Transmission of 2019-ncov infection from an asymptomatic contact in germany. New England Journal of Medicine, 2020.

[29] Richard D Smith. Responding to global infectious disease outbreaks: lessons from sars on the role of risk perception, communication and management. Social science & medicine, 63(12):3113–3123, 2006.

[30] Peter X-K. Song. Correlated Data Analysis: Modeling, Analytics, and Applications. Springer, 2007.

[31] Peter Xue-Kun Song. Monte carlo kalman filter and smoothing for multivariate discrete state space models. Canadian Journal of Statistics, 28(3):641–652, 2000.

[32] Lorenzo Subissi, Clara C Posthuma, Axelle Collet, Jessika C Zevenhoven-Dobbe, Alexander E Gorbalenya, Etienne Decroly, Eric J Snijder, Bruno Canard, and Isabelle Imbert. One severe acute respiratory syndrome coronavirus protein complex integrates processive rna polymerase and exonuclease activities. Proceedings of the National Academy of Sciences, 111(37):E3900–E3909, 2014.

[33] Haoxuan Sun, Yumou Qiu, Han Yan, Yaxuan Huang, Yuru Zhu, and Song Xi Chen. Tracking and predicting covid-19 epidemic in china mainland. medRxiv, 2020.

[34] Chen Wang, Peter W Horby, Frederick G Hayden, and George F Gao. A novel coronavirus outbreak of global health concern. The Lancet, 2020.

[35] WHO. Emergencies preparedness, response. pneumonia of unknown origin – china. disease outbreak news. Available online: https://www.who.int/csr/don/12-january-2020-novel-coronavirus-china/en/ accessed on 05 February 2020, 2020.

[36] Hadley Wickham. ggplot2: Elegant Graphics for Data Analysis. Springer-Verlag New York, 2016.

[37] Yu-Tao Xiang, Wen Li, Qinge Zhang, Yu Jin, Wen-Wang Rao, Liang-Nan Zeng, Grace KI Lok, Ines HI Chow, Teris Cheung, and Brian J Hall. Timely research papers about covid-19 in china. The Lancet, 2020.

[38] Xiao-Wei Xu, Xiao-Xin Wu, Xian-Gao Jiang, Kai-Jin Xu, Ling-Jun Ying, Chun-Lian Ma, Shi-Bo Li, Hua-Ying Wang, Sheng Zhang, Hai-Nv Gao, et al. Clinical findings in a group of patients infected with the 2019 novel coronavirus (sars-cov-2) outside of wuhan, china: retrospective case series. BMJ, 368, 2020.

[39] Guangchuang Yu. nCov2019: Stats of the ‘2019-nCov’ Cases, 2020. R package version 0.0.8.

[40] Sheng Zhang, MengYuan Diao, Wenbo Yu, Lei Pei, Zhaofen Lin, and Dechang Chen. Estimation of the reproductive number of novel coronavirus (covid-19) and the probable outbreak size on the diamond princess cruise ship: A data-driven analysis. International Journal of Infectious Diseases, 2020.

[41] Bin Zhu, Jeremy MG Taylor, and Peter X-K Song. Signal extraction and breakpoint identification for array cgh data using robust state space model. arXiv preprint 1201.5169, 2012.

[42] Na Zhu, Dingyu Zhang, Wenling Wang, Xingwang Li, Bo Yang, Jingdong Song, Xiang Zhao, Baoying Huang, Weifeng Shi, Roujian Lu, et al. A novel coronavirus from patients with pneumonia in china, 2019. New England Journal of Medicine, 2020.

